# Estimating the reproduction number and transmission heterogeneity from the size distribution of clusters of identical pathogen sequences

**DOI:** 10.1101/2023.04.05.23287263

**Authors:** Cécile Tran-Kiem, Trevor Bedford

## Abstract

Quantifying transmission intensity and heterogeneity is crucial to ascertain the threat posed by infectious diseases and inform the design of interventions. Methods that jointly estimate the reproduction number *R* and the dispersion parameter *k* have however mainly remained limited to the analysis of epidemiological clusters or contact tracing data, whose collection often proves difficult. Here, we show that clusters of identical sequences are imprinted by the pathogen offspring distribution, and we derive an analytical formula for the distribution of the size of these clusters. We develop and evaluate a novel inference framework to jointly estimate the reproduction number and the dispersion parameter from the size distribution of clusters of identical sequences. We then illustrate its application across a range of epidemiological situations. Finally, we develop a hypothesis testing framework relying on clusters of identical sequences to determine whether a given pathogen genetic subpopulation is associated with increased or reduced transmissibility. Our work provides new tools to estimate the reproduction number and transmission heterogeneity from pathogen sequences without building a phylogenetic tree, thus making it easily scalable to large pathogen genome datasets.

**Significance statement:** For many infectious diseases, a small fraction of individuals has been documented to disproportionately contribute to onward spread. Characterizing the extent of superspreading is a crucial step towards the implementation of efficient interventions. Despite its epidemiological relevance, it remains difficult to quantify transmission heterogeneity. Here, we present a novel inference framework harnessing the size of clusters of identical pathogen sequences to estimate the reproduction number and the dispersion parameter. We also show that the size of these clusters can be used to estimate the transmission advantage of a pathogen genetic variant. This work provides crucial new tools to better characterize the spread of pathogens and evaluate their control.

## Introduction

Characterizing transmission parameters describing the intensity and heterogeneity of infectious diseases’ spread is fundamental both to understand the threat posed by epidemics and to inform their control. The reproduction number *R*, which describes the average number of secondary infections engendered by a single primary case, is a key statistic as it directly reflects the ability for a pathogen to propagate within a population (corresponding to *R > 1*) (1–4). Additionally, heterogeneity in the contribution of individuals to disease spread has important implications for outbreak control. When a small proportion of individuals plays a disproportionate role in transmission, control strategies targeting these *superspreaders* can largely reduce the epidemic burden with a small effort (1,5). Individual variation is typically measured with the dispersion parameter *k*, with lower values corresponding to a higher degree of heterogeneity (5).

Estimates of *R* can be obtained from the analysis of epidemiological time series (*e.g.* cases, hospitalizations or deaths). These methods have widely been used to track in real-time changes in transmission rates or the impact of interventions (2,6,7). However, these approaches do not allow the estimation of the dispersion parameter *k*. Other methods have been proposed to jointly infer the reproduction number and the dispersion parameter from transmission chain data or epidemiological cluster sizes (5,8–10). The collection of such data relies on the identification of epidemiological relationships between infected individuals. This can however prove difficult if there is widespread community transmission (hindering our ability to identify the putative infector), if a fraction of infected individuals remains undetected or if the resources available for such investigation remain scarce.

Pathogen genome sequences can provide insights into the proximity of individuals in a transmission chain (11–13) or into the epidemiological processes associated with their spread (14). Phylodynamic approaches have been widely used to estimate epidemiological parameters from the shape of phylogenies, including the reproduction number *R* (15–18). Current methods however face a number of limitations. First, they require intensive computation and therefore do not scale well to large datasets. New approaches to estimate transmission parameters from sequence data without resorting to subsampling are hence of primary interest. Second, they generally do not enable to estimate the extent of transmission heterogeneity (superspreading or *k*). Finally, polytomy-rich phylogenies tend to decrease the statistical power available to estimate growth rates, thus deprecating the value of these methods during the early stage of an outbreak or when large superspreading events occur.

Here, we demonstrate that the size distribution of clusters of identical sequences (or of polytomies) is shaped by epidemic transmission parameters (*R* and *k*). We develop and evaluate a statistical model accounting for heterogeneity in transmission to estimate the reproduction number *R* and the dispersion parameter *k* from this distribution. Our method does not require building the associated phylogenetic tree and is suited for the analysis of sequence data from pathogens causing acute infections with narrow transmission bottlenecks. Applying our framework to different epidemiological situations, we recover expected transmission parameters for well-characterized pathogens. Finally, we develop a novel inference framework to quantify differences in the transmissibility of genetic variants.

## Results

### Distribution of the number of offspring with identical genomes

In this work, we used the formalism introduced by Lloyd-Smith et al. (5) and assumed that the number of secondary cases (or offspring) generated by a single index case follows a negative binomial distribution of mean *R* (the reproduction number) and dispersion parameter *k*. We focused here on the spread of a pathogen that mutates. During replication within a host, pathogens accrue mutations, generating some level of within-host diversity, that can be passed to an infectee and in which they can reach fixation. To account for this, we are looking at consensus sequences, which are representative genetic sequences composed of the most frequent nucleotides at each site. We use the term *substitution* to refer to differences in pathogen sequences at the consensus level. Here, we were interested in the number of offspring which have the same consensus genome sequence as their infector (Figure 1A) (19). We introduced *p*, the probability that an infectee has the same consensus genome as their infector. We showed that the number of offspring with identical genomes follows a negative binomial distribution of mean *p R* and dispersion parameter *k* (see Materials and methods). This highlights that the relative timescale at which substitution and transmission events typically occur (captured by *p*) along with transmission intensity (captured by R), will shape the expected mean number of secondary cases with identical sequences (Figure 1B).

**Figure 1:**
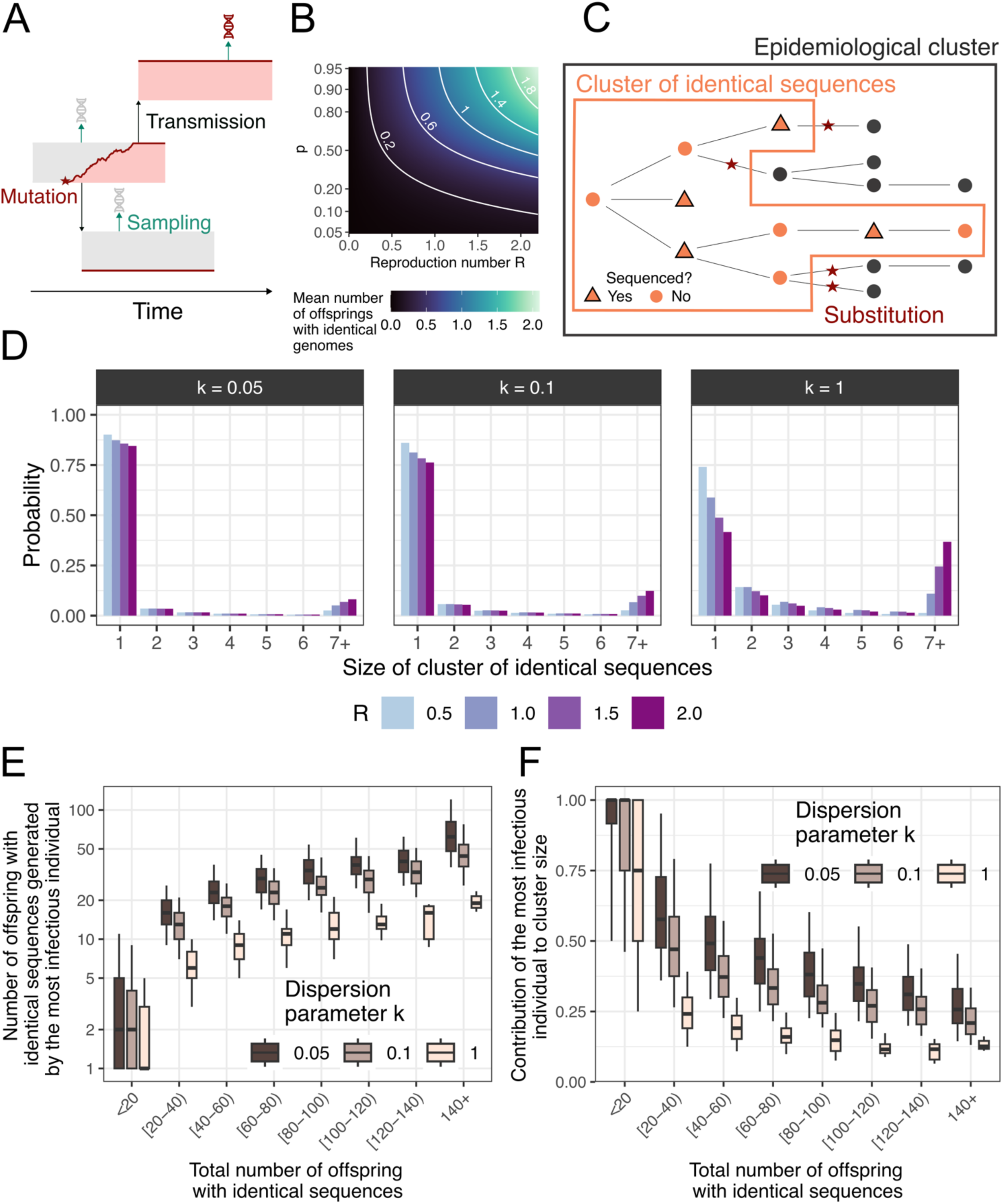
Impact of the reproduction number *R* and the dispersion parameter *k* on the size distribution of clusters of identical sequences. **A.** Mutations occur within infected host and might result in infector-infectee pairs having different consensus sequences. **B.** Mean number of offspring with identical genomes as a function of the reproduction number and the probability *p* that an infector and an infectee have the same consensus sequence (logit scale for the y-axis). **C.** Relationship between epidemiological clusters and clusters of identical sequences. **D.** Probability mass function of the size of clusters of identical sequences for different values of *R* and *k* and assuming a probability that an infector and infectee have the same consensus sequence of 0.7. **E.** Number of offspring with identical sequences generated during the largest transmission event as a function of the total number of offspring with identical genomes generated by a primary case exploring different values for *k*. **F.** Contribution of the largest transmission event to the total number of offspring with identical sequences as a function of the total number of offspring with identical genomes generated by a primary case exploring different values for *k*. In E-F, we report for each value of *k* the results of 100,000 simulated clusters of identical sequences with a reproduction number *R* of 1 and a probability that an infector and an infectee have the same consensus sequence of 0.7. In E-F, the boxplots indicate the median along the 5%, 25%, 75% and 95% quantiles. In E-F, the total number of offspring with identical sequences is equal to the size of a cluster of identical sequences minus 1.

### Size of clusters of identical sequences

We defined clusters of identical sequences as subsets of epidemiological clusters, which are defined as groups of infections with a known epidemiological link (9). Whereas epidemiological clusters end when every transmission chain composing them ceases to circulate (*i.e.* does not produce any offspring), we define clusters of identical sequences as ending when each transmission chain dies out or results in a substitution event (Figure 1C). Figure 1D depicts how the distribution of the size of clusters of identical sequences was impacted by assumptions regarding the transmission parameters *R* and *k*. For example, higher values of R would result in larger clusters of identical sequences. For a fixed *R*, lower values of *k* (corresponding to a higher heterogeneity in transmission) would result in a lower probability for a cluster to reach a certain size.

### Contribution of highly infectious individuals to the size of polytomies

Though a highly infectious individual (who generates a large number of offspring) will tend to generate larger clusters of identical sequences, the presence of large polytomies in a phylogeny is not necessarily the signature of a high degree of transmission heterogeneity. When the mean number of offspring with identical genomes (equal to *R* ⋅ *p*) is greater than 1, the probability of cluster extinction is strictly lower than 1 (Figure S1). Large clusters of identical sequences are thus not unlikely when the reproduction number *R* is greater than 1/*p*, regardless of the extent of transmission heterogeneity (Figure 1D, S1). This value corresponds to the criticality threshold for transmission events associated with identical sequences. Moreover, large clusters of identical sequences do not stem solely from a single transmission generation as they also encompass earlier or later transmission events. In Figure 1E-F, we explored to what extent the most infectious individual of a cluster was contributing to the size of that cluster. We found that larger clusters of identical sequences corresponded to more infectious individuals (Figure 1E). Assuming a reproduction number *R* of 1 and a probability p that an infector and an infectee have the same consensus sequence of 1, the median contribution of the most infectious individual (defined as the cluster member generating the most offspring) to overall cluster size was however limited for very large clusters of identical sequences (e.g. 13%, 17% and 21% for values of *k* of 1.0, 0.1 and 0.05 considering clusters of identical sequences greater than 140).

### Inference of transmission parameters from the size distribution clusters of identical sequences

Clusters of identical sequences are hence imprinted by the characteristics of the disease offspring distribution. This suggests that the size distribution of these clusters might be used to estimate outbreak transmission parameters. We thus next sought to develop an inference framework to estimate the reproduction number *R* and the dispersion parameter *k* from the size distribution of clusters of identical sequences. Our maximum-likelihood based approach was inspired by the methods used to estimate these two parameters from the size distribution of epidemiological clusters (8,9). It requires input values for the probability p that an infector and an infectee have the same consensus sequence and the fraction of infections that would be sequenced.

We evaluated our inference framework on simulated data generated under different assumptions regarding the reproduction number, the dispersion parameter, the probability that an infector and an infectee have the same consensus sequence and the fraction of infections sequenced. Figure 2A depicts the relationship between estimates of the reproduction number *R* and the true value used to generate synthetic clusters. We were able to correctly estimate the reproduction number at lower values. However, when the reproduction number reached the threshold of 1/*p*, we became unable to accurately estimate its value, with our estimates remaining *stuck* at the threshold value. Below the reproduction number threshold of 1/*p*, increasing the number of clusters included in our analysis improved the precision of our estimates (Figure S2). When *R* remained below the threshold, we were able to correctly infer the value of the dispersion parameter *k* (Figure 2B, Figure S3). Relying on a low number of clusters for the inference resulted in overestimates of the dispersion parameter (Figure S3) and underestimates of the reproduction number (Figure S2), in line with previous studies on the ability to infer parameters from a negative binomial distribution (9,20). For values of *R* higher than the threshold, we were unable to correctly infer the dispersion parameter, with values consistently overestimated and a bias that increased with both k and the expected mean number of offspring of identical sequences (Figure S4-S5). Considering lower values for the proportion of infections sequenced required a greater number of clusters to be analyzed to reach the same accuracy as that obtained at higher sequencing fraction (Figure S6). Overall, we found that bias increased with the proportion of singletons among clusters (Figures S7).

**Figure 2:**
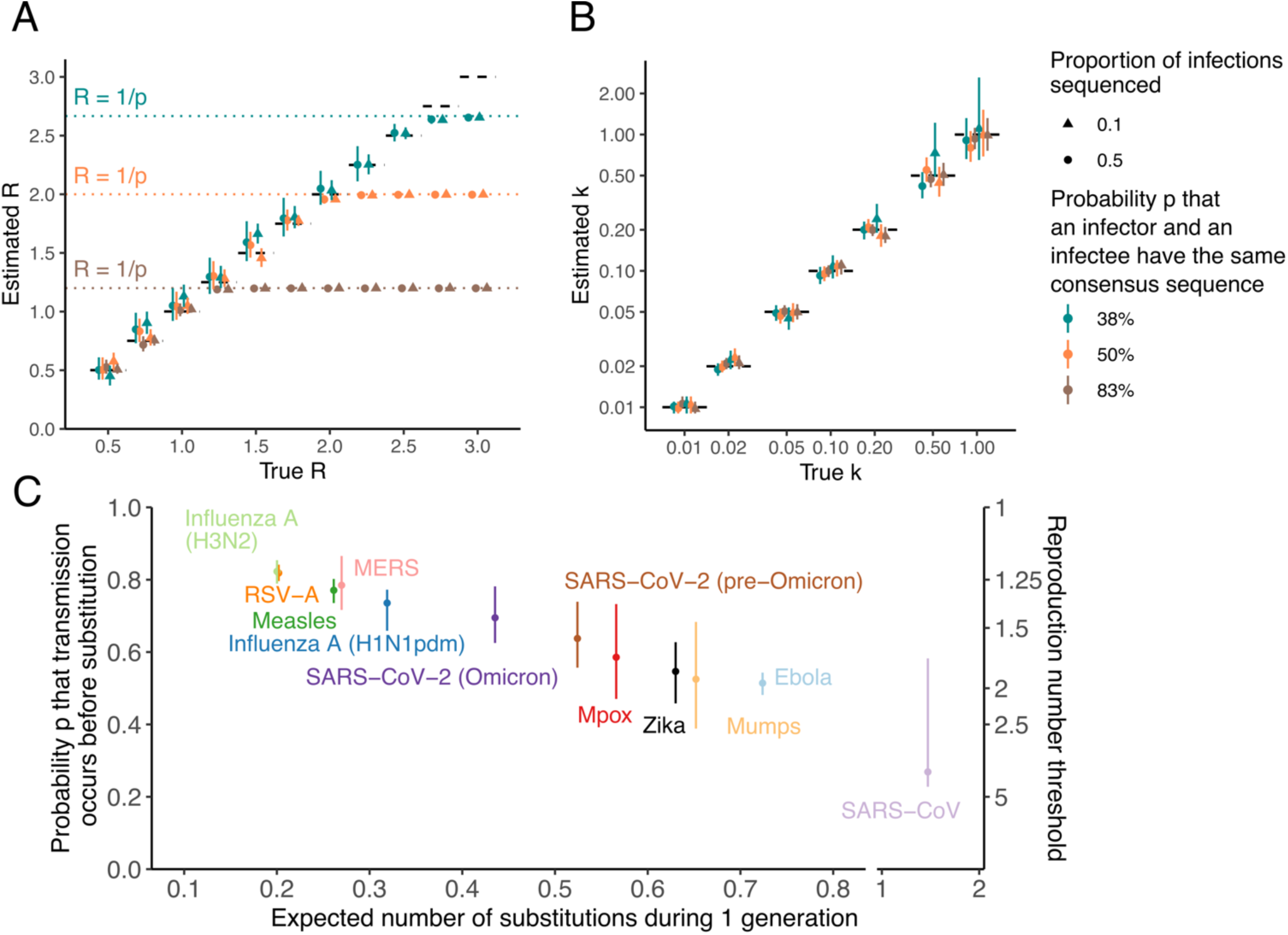
Inference of the reproduction number and the dispersion parameter from synthetic clusters of identical pathogen sequences. **A.** Estimated reproduction number *R* as a function of the true reproduction number value used to simulate synthetic clusters. **B.** Estimated dispersion parameter *k* as a function of the true dispersion parameter value used to simulate synthetic clusters. **C.** Estimates of the probability *p* that transmission occurs before substitution for a range of pathogens. In C, points indicate central estimates and vertical segments indicate uncertainty ranges (see methods). In A and B, point estimates correspond to maximum-likelihood estimates and vertical segments to 95% likelihood profile confidence interval obtained from analyzing 1000 synthetic clusters of identical sequences. Results in A correspond to the joint inference of *R* and *k* for true values of k equal to 0.1. Results in B correspond to the joint inference of *R* and *k* for true values of *R* equal to 1.0. Different proportions of sequencing among infections are explored (10% and 50%). In A and B, different probabilities *p* that an infector and an infectee have the same consensus sequence are explored.

### Reproduction number threshold for different acute infections

The *R* threshold value below which our inference framework will produce unbiased estimates will thus depend both on the natural history of the infection and the evolutionary characteristics of the pathogen. We explored how this threshold varied for a range of viruses (Figure 2C, Table S1-S2). This required estimating the probability *p* that an infector and an infectee have the same consensus sequence. For pathogens causing acute infections characterized by a narrow transmission bottleneck, we approximated *p* by the probability that a transmission event occurs before a substitution event (Supplementary text A). For Severe Acute Respiratory Syndrome (SARS), we estimated a probability for transmission to occur before substitution of 27% (uncertainty range: 23%-58%), thus resulting in a R threshold value higher than 3. This observation is consistent with previous work highlighting the value of genome sequences in reconstructing SARS-CoV-like outbreaks (11). For the other pathogens we considered, we estimated values of the probability that transmission occurs before substitution between 51% (48%-54%) for Ebola and 82% (79%-85%) for H3N2 influenza, corresponding to *R* thresholds between 1.22 (1.18-1.26) and 1.96 (1.85-2.08).

Our inference framework hence provides unbiased estimates of both *R* and *k* when the mean expected number of offspring with identical genomes lies below 1. When reaching this critical threshold, estimates of the reproduction number become biased downwards. In the following, we focus on situations where the reproduction number lies below the critical threshold of 1/*p* and we report a series of analyses showcasing how our method may be applied in different epidemiological settings. For each of these analyses, the fraction of infections sequenced was computed as the product of the proportion of cases sequenced and the assumed proportion of infections detected as cases.

### Recovering characteristics of the Middle East Respiratory Syndrome (MERS) outbreak (2013–2015)

MERS is a respiratory infection first identified in 2012, associated with a case fatality ratio of around 40%. MERS is transmitted to humans either upon contact with infected camels, who act as a zoonotic reservoir, or with infected humans (21). Human-to-human transmission however results in subcritical transmission chains (*R < 1*) (22–26) characterized by substantial heterogeneity (24,25). We analyzed 174 MERS-CoV human sequences sampled between 2013 and 2015 (26) and identified 140 clusters of identical sequences, with an average cluster size of 1.2 (Figure 3A). Assuming all infections were detected as cases, we estimated a reproduction number of 0.57 (95% confidence interval (CI): 0.46-0.70) and a dispersion parameter of 0.14 (95% CI: 0.04-0.46) (Figure 3B-C, Table S3). These estimates were consistent with those obtained from the analysis of the size of MERS epidemiological clusters (24). Interestingly, our confidence intervals were narrower than the ones obtained from the analysis of epidemiological clusters.

**Figure 3:**
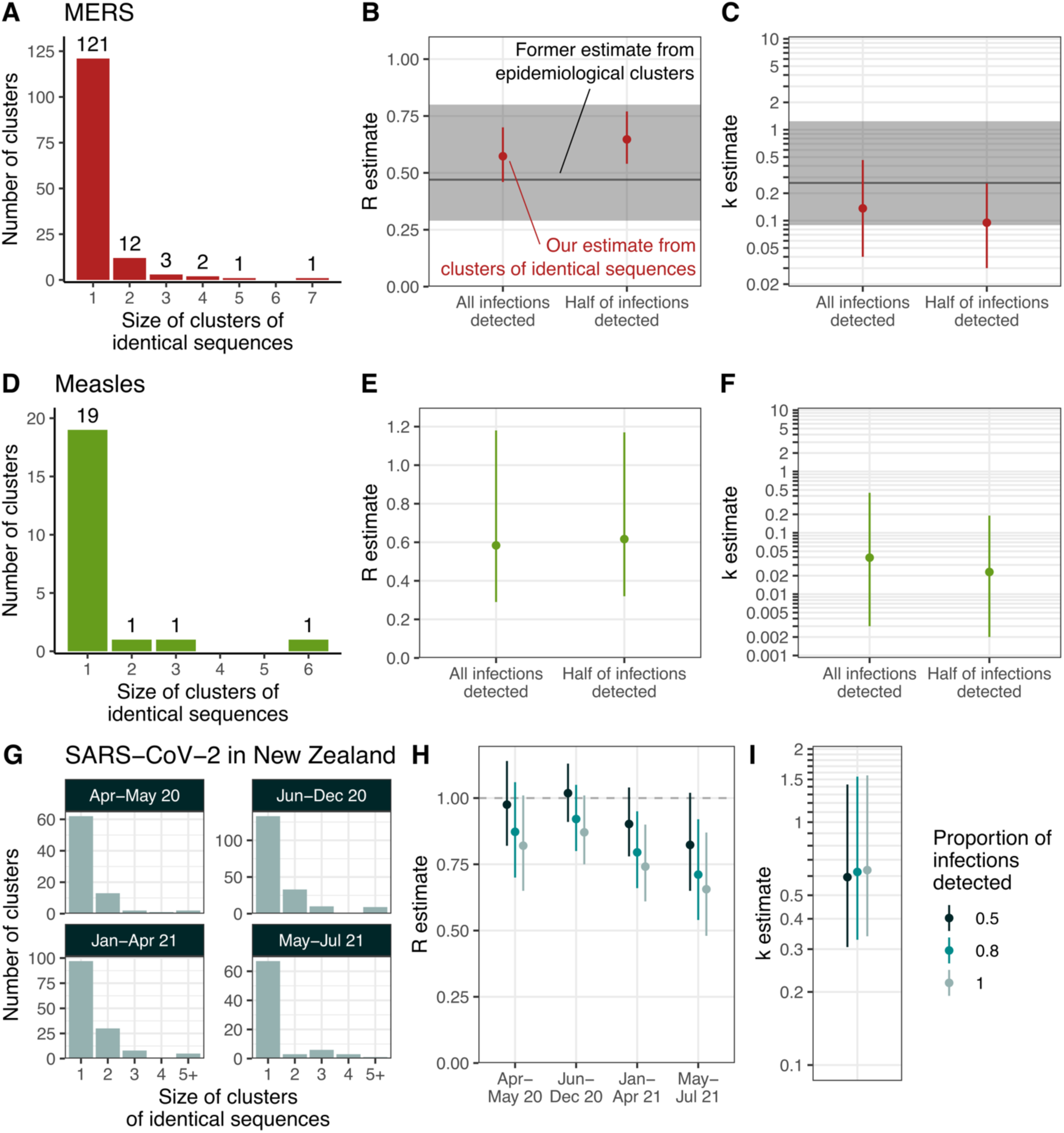
Application of our modelling framework to different pathogen genome datasets. **A.** Distribution of the size of clusters of identical sequences during the 2013-2015 MERS outbreak (26). Estimates for MERS of **B.** the reproduction number *R* and **C.** the dispersion parameter *k* assuming that all or half of infections were detected as cases. **D.** Distribution of the size of clusters of identical sequences during the measles 2017 outbreak in the Veneto region in Italy (30). Estimates for measles of **E.** the reproduction number *R* and of **F.** the dispersion parameter *k* assuming that all or half of infections were detected as cases. **G.** Distribution of the size of clusters of identical sequences during the COVID- 19 pandemic in New Zealand by period. Estimates of **H.** the reproduction number *R* and **I.** the dispersion parameter *k* for SARS-CoV-2 in New Zealand. For B, C, E, F, H and I, the points correspond to maximum likelihood estimates and the vertical segments to 95% likelihood profile confidence intervals. In B and C, the horizontal lines (respectively the shaded area) correspond to the maximum-likelihood estimate (respectively the 95% confidence intervals) obtained by Kucharski and Althaus (24) from the analysis of the size of MERS epidemiological clusters.

Epidemiological surveillance is generally able to only detect a fraction of the overall infection burden and is skewed towards symptomatic or severe outcomes. For MERS, modelling studies have suggested that detected cases may have accounted for only around half of overall infections (27). In this scenario, we estimated a higher value of 0.65 (95%CI: 0.54-0.77) for the reproduction number and a lower value of 0.09 (95%CI: 0.03-0.26) for the dispersion parameter (corresponding to a higher degree of heterogeneity in transmission) (Figure 3B-C, Table S3). Estimates were however qualitatively similar to the ones obtained assuming infections were completely detected.

### Characterizing measles transmission in the post-vaccination era

During the last decade, European countries have experienced important measles outbreaks despite elevated vaccination coverages. Such outbreaks may occur either due to insufficient population vaccination coverage or due to persisting pockets of low vaccination rates (28,29). Here, we estimated transmission parameters during the 2017 measles outbreak that occurred in the Veneto Region in Italy from 30 sequences sampled during this time period (30) (Figure 3D). Assuming all infections were detected as cases, we estimated a reproduction number *R* of 0.58 (95%CI: 0.29–1.18) and a dispersion parameter *k* of 0.04 (95%CI: 0.003-0.45) (Figure 3E-F, Table S4). We also conducted a sensitivity analysis assuming that 50% of infections may have gone undetected. We obtained an estimate of *R* of 0.62 (95%CI: 0.32-1.17) and of *k* of 0.02 (95%CI: 0.002-0.19) (Figure 3E-F, Table S4). Our estimates of the reproduction number were similar to those obtained for measles outbreaks occurring in high-income countries in the post-vaccination era (10,31). Due to the limited number of clusters of identical sequences included in our analysis, the uncertainty around our estimate of the dispersion parameters remained quite substantial. Our results yet suggest a high degree of heterogeneity, in line with previous analyses (5,10).

### Severe acute respiratory syndrome coronavirus 2 (SARS-CoV-2) transmission heterogeneity during a Zero-COVID strategy

We next focused on the characteristics of the coronavirus disease 19 (COVID-19) epidemic in New Zealand between April 2020 and July 2021, a period during which the epidemic was mostly suppressed without reported community transmission. We applied our modelling framework to SARS-CoV-2 sequences collected during this timeframe. We split the study period into 4 subperiods: April-May 2020, June-December 2020, January-April 2021 and May-July 2021 (Figure 3G) during which respectively 25%, 51%, 46% and 48% of cases were sequenced. We allowed the reproduction number to vary between periods but assumed that transmission heterogeneity remained constant throughout. Assuming that 80% of infections were detected as cases, we estimated reproduction numbers below unity throughout the period (Figure 3H, Table S5) and a dispersion parameter *k* of 0.63 (95%CI: 0.34-1.56) (Figure 3I, Table S5), which corresponds to 23-25% (17-33%) of individuals being responsible for 80% of infections throughout the period. We explored the impact of our assumption regarding the fraction of infections detected as cases on our estimates and found that values ranging between 50 and 100% would not quantitatively change our findings (Figure 3H-I, Table S5). Our results are consistent with previous SARS-CoV-2 overdispersion estimates (32,33).

In these 3 case studies, estimates of the reproduction number were little impacted by uncertainty around *p* (Figures S8-S9, Tables S3-S7). Estimates of the dispersion parameter were almost not impacted by uncertainty around *p*.

### Monitoring the transmission advantages of genetic variants

In the previous sections, we looked at how we can use the size distribution of polytomies to estimate *R* and *k*. We now focused on how the spread of a variant characterized by a transmission advantage can influence the size distribution of clusters of identical sequences. More specifically, we were interested in whether hosts infected with a certain genetic subpopulation significantly infect more individuals than hosts infected with another genetic subpopulation. We aimed to quantify this transmission advantage, which may be impacted by the intrinsic transmissibility of the variant of interest, its ability to escape pre-existing immunity or certain characteristics of the host population (*e.g.* immunity profiles). We developed an inference framework based on statistical hypothesis testing (see Methods) to determine whether some variant and non-variant pathogens are associated with different reproduction numbers from the size distribution of clusters of identical sequences observed in these two genetic populations (Figure 4A). In the following, we will refer to variant as the more transmissible genetic subpopulation. However, the role of the variant and the non-variant are interchangeable, making our results hence directly applicable for a less transmissible one.

**Figure 4:**
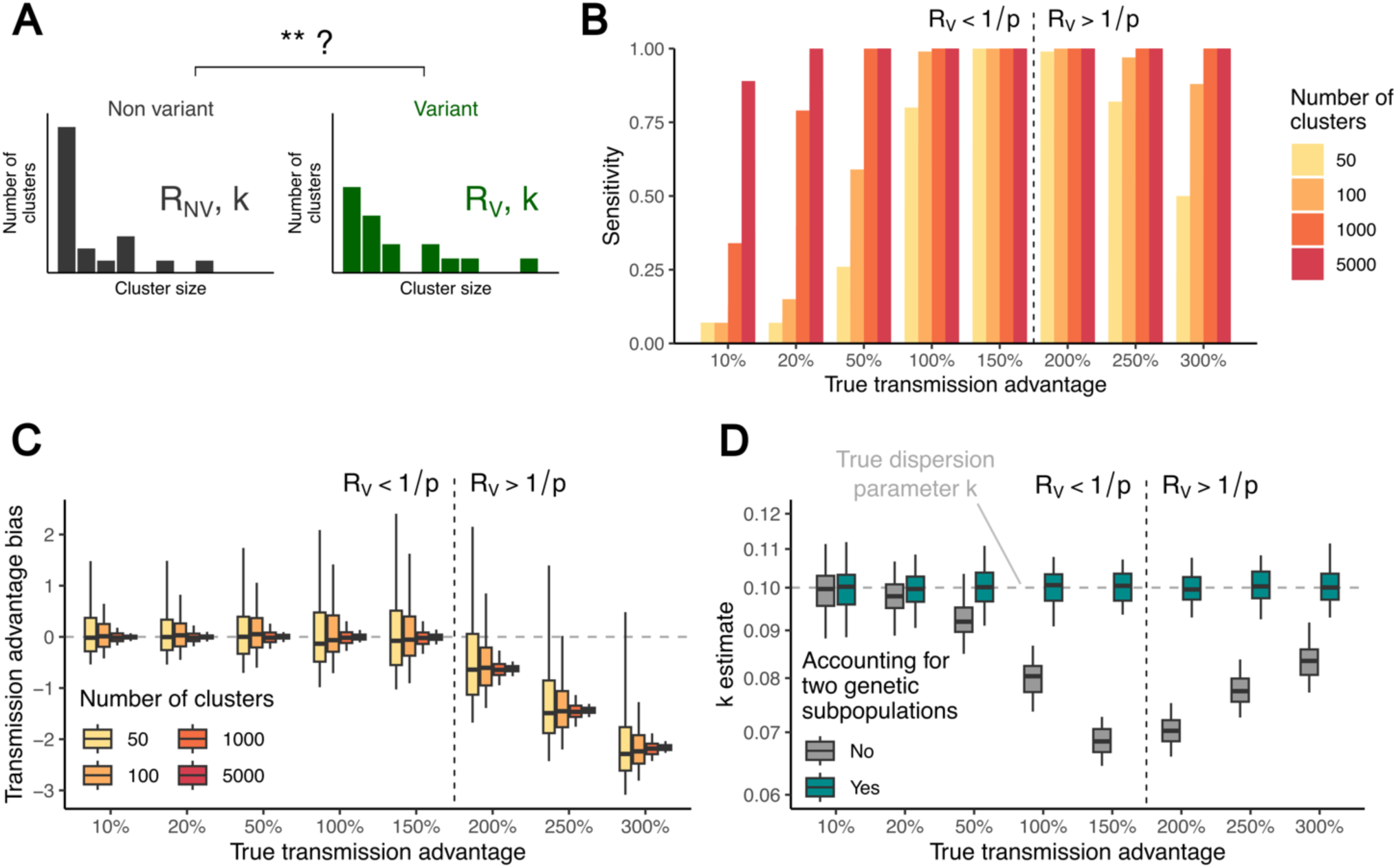
Performance of our inference framework in quantifying the transmission advantage of genetic variants from the size distribution of polytomies. **A.** We aim to compare the size distribution of clusters of identical sequences splitting a given pathogen between variant and non-variant subtypes. **B.** Sensitivity of the analysis of clusters of identical pathogen sequences in detecting a change in the transmissibility of a genetic variant. This is done for different values of the true transmission advantage and exploring different sizes of datasets on which the inference is based. **C.** Transmission advantage bias for different values of the true transmission advantage and exploring different sizes of datasets on which the inference is based. **D.** Impact of accounting for two genetic subpopulations on estimates of the dispersion parameter *k*. The transmission advantage bias is defined as 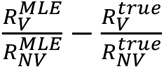 where 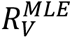 (respectively 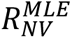) is the maximum likelihood estimate for the reproduction number of the variant (respectively the non-variant) and 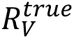 (respectively 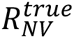) is the true reproduction number of the variant (respectively the non-variant) used to generate synthetic cluster data. In B-D, the vertical dashed line delimits the regions where the reproduction number of the variant is above/below the threshold of *1/p*. In B-D, the results correspond to scenarios where the true dispersion parameter is equal to 0.1, the probability that an infector and an infectee have the same consensus sequence to 0.5 and the reproduction number of the non-variant to 0.75. In D, the results correspond to analyses based on 5,000 clusters of identical sequences for both the variant and the non-variant.

We evaluated the ability of our statistical framework to detect the presence of a transmission advantage given the number of genetic clusters observed and the magnitude of this transmission advantage (Figure 4B). We found that (i) the sensitivity of our test increased when more clusters of identical sequences were observed and that (ii) the detection of small transmission advantages required the analysis of a larger number of clusters. When the reproduction number of the variant reached the threshold of 1/*p*, the sensitivity of our test decreased. We then evaluated the ability of our framework to estimate the variant transmission advantage. We found that we have unbiased estimates of the transmission advantage as long as both the reproduction number of the variant and the non-variant remained below the 1/*p* threshold (Figure 4C, Figure S10). Analyzing a greater number of clusters increased the precision of our test (Figure 4C). Interestingly, sensitivity remained high even above the threshold of 1/*p* when a sufficiently large number of clusters were included in the analysis. We also found that failing to allow for the reproduction number to differ between two genetic subpopulations resulted in overestimating the extent of transmission heterogeneity (Figure 4D, Figure S11).

### Application to SARS-CoV-2 variants in Washington state

To illustrate our framework, we analyzed SARS-CoV-2 sequence data collected in Washington state, in the United States (Figure 5A, Figure S12). Figure 5B depicts the mean size of clusters of identical sequences for different variants depending on the month of first detection of the cluster in Washington state. We found that the dynamics of the mean size of clusters of identical sequences reflected the dynamics of strain replacement in Washington state (Figure 5A), with greater average cluster sizes being observed when a given variant of concern was increasing in proportion. This pattern is expected as we showed that average cluster sizes can be related to the effective reproduction number. Other elements (such as the fraction of infections sequenced) can impact patterns of cluster sizes.

**Figure 5:**
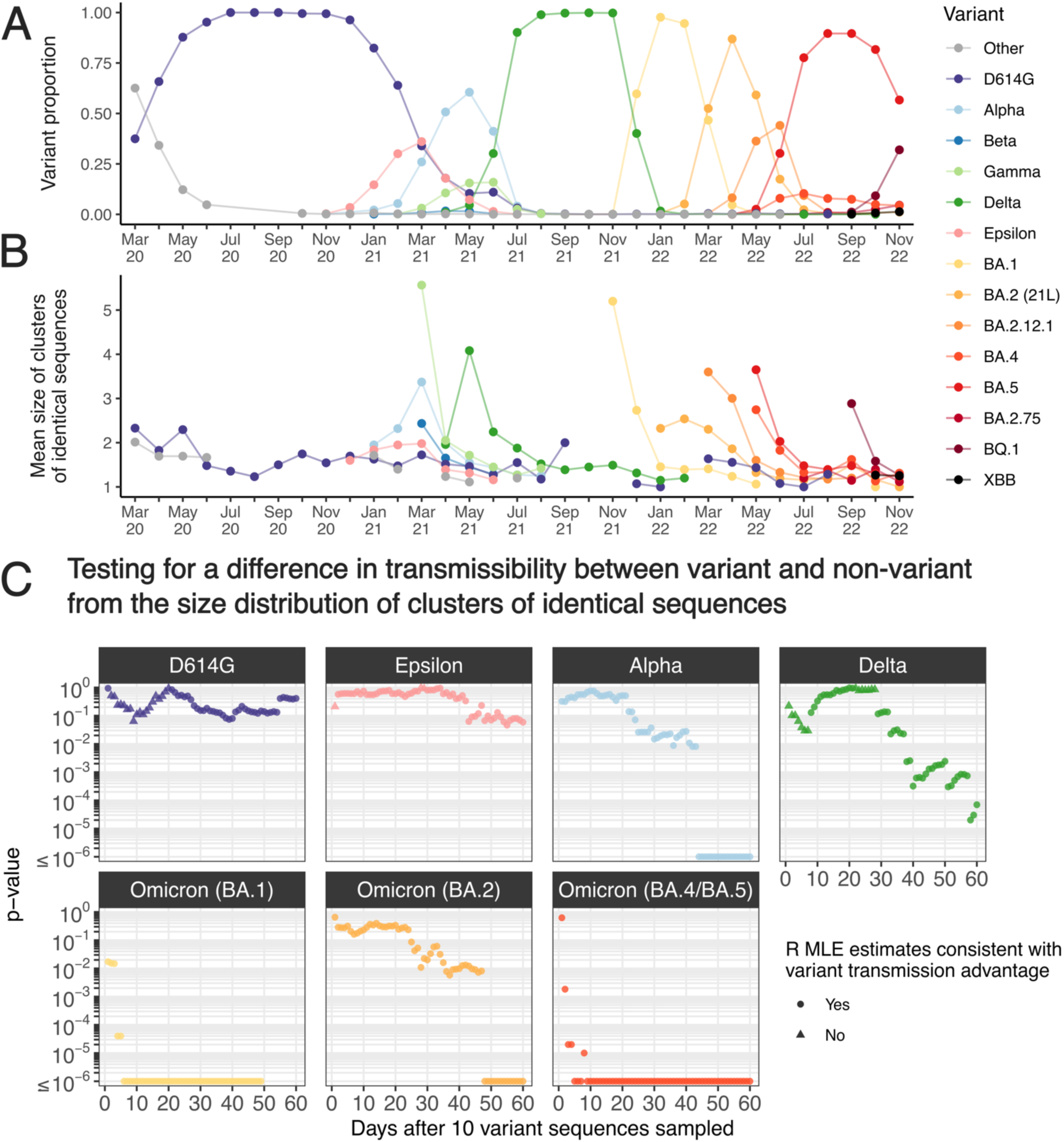
Analysis of SARS-CoV-2 clusters of identical sequences in Washington state. **A.** Proportion of variant among sequences by month of sample collection. **B.** Mean size of clusters of identical sequences in Washington state from March 2020 across different SARS-CoV-2 clades. **C.** P- values over time since collection of 10 variant sequences for different SARS-CoV-2 variants during the COVID-19 pandemic in Washington state assuming 50% of infections were detected as cases. In B, we report the mean size of clusters of identical sequences grouping clusters depending on their first detection date. In B, we display the mean cluster size for a given month and a given variant if at least five clusters of identical sequences were identified (for this variant this month). The variants dynamics were derived based on Nextstrain clades allocation (34,35). In C, we considered maximum likelihood estimated (MLE) to be consistent with a variant transmission advantage if the estimated reproduction number of the variant was higher than that of the non-variant.

In the early stages of their spread, variants of concerns (VOCs) such as Alpha, Delta or Omicron have been associated with initially short doubling times (36–39), corresponding to reproduction numbers above the threshold of 1/*p*. This, alongside other factors such as changes in the reproduction number over time or the implementation of stringent control measures to slow the epidemic spread, could result in biased estimates of the transmission advantage. To overcome this caveat and avoid overinterpreting our results, we instead focused on whether we were able to detect a transmission change as we previously saw that sensitivity remained high for large cluster datasets. We tested a list of variants on whether they were associated with a transmission advantage (or disadvantage) compared to the other strains that were circulating at that time. For each variant, we evaluated how our conclusions were impacted by the width of the analysis window. In Figure 5C, we depicted the p-values for different variants as a function of the time before which clusters had the be first observed to be included in the analysis (40,41). This was done assuming that 50% of infections were detected as cases. For Alpha, Delta, and Omicron (BA.1, BA.2 and BA.4/BA.5), we found statistical support for their increased transmissibility compared to other lineages that circulated at that time. Across variants, p-values decreased with increasing time window length. For the Omicron sub-variants BA.1 and BA.4/BA.5, we found a strong statistical support (p-value < 10^-4^) for an increased transmissibility considering time-windows as short as 10 days. We did not find evidence for the D614G mutation, that occurred early on during the pandemic, to be associated with an increased transmissibility, which has been suggested by other studies (40,41). For the Epsilon VOC, the lowest p-value we found equaled 0.046. This suggests that it might be associated with an increased transmission intensity. We obtained similar conclusions when varying our assumption regarding the fraction of infections detected as cases (Figure S13).

## Discussion

The extent of heterogeneity in transmission and the transmissibility of a pathogen have important implications regarding both its potential epidemic burden and the impact of control measures. Despite their epidemiological relevance, estimating these two parameters has remained delicate during most outbreaks. In this work, we presented a novel modelling framework enabling the joint inference of the reproduction number *R* and the dispersion parameter *k* from the size distribution of clusters of identical sequences. We evaluated the performance of our statistical framework and subsequently applied it to a range of epidemiological situations. Finally, we showed how it may be extended to look at the relative transmissibility of different genetic subpopulations.

Robust estimates of key epidemiological parameters (such as the reproduction number *R* and the dispersion parameter *k*) are critical to ascertain epidemic risks. They can be obtained by directly analyzing chains of transmission (5), though such data are generally difficult or almost impossible to collect for some pathogens (42). Establishing epidemiological links between cases may indeed be hampered by widespread community transmission, sub-clinical disease manifestation or when the infection is spread through a vector. Alternatively, it has been shown that the size distribution of epidemiological clusters can be harnessed to obtain such estimates (9,43). Beyond the potential challenges in establishing epidemiological links between cases, apparent epidemiological clusters may involve different transmission chains, that couldn’t be disentangled without further molecular investigation. This is for example the case in the measles outbreak reported in Pacenti et al. (30), that we analyzed here, where one observed epidemiological cluster was actually constituted of two evolutionary groups. Relying on the size of epidemiological clusters would thus likely lead to overestimating the reproduction number. Here, we proposed a new approach, exploiting the relationship between epidemiological clusters and clusters of identical sequences to provide robust estimates of both *R* and *k* which does not require to reconstruct these epidemiological relationships. Interestingly, we obtained narrower confidence intervals for MERS transmission parameters than the ones obtained from the analysis of the size of epidemiological clusters (24). This can likely be explained by the greater number of clusters (though smaller) included in our analysis. This suggests that even in settings where transmission chains may be reconstructed, the combination of case investigation with pathogen sequencing may be valuable to increase statistical power and our ability to estimate these key epidemiological parameters. Moreover, as estimates may be biased upwards for the dispersion parameter and downwards for the reproduction number when relying on a low number of clusters for the analysis (Figure S2-S3) (9,20), the inclusion of a larger number of clusters enabled by looking at genetic clusters instead of epidemiological ones can reduce biases.

High-throughput sequencing has enabled the faster and cheaper generation of pathogen genome sequences. Current tree-based methods nonetheless require heavy computations to estimate growth rates from pathogen genomes. Here, we showed that the size of clusters of identical sequences directly contains an imprint of the underlying epidemiological processes and can be leveraged to characterize the intensity and heterogeneity in transmission, this without requiring the estimation of the associated phylogenetic tree. Moreover, the speed and ease of implementation of our method could make it valuable for public health professionals who may not have access to or be comfortable using scientific software programs currently used for phylodynamic analysis. The concept of phylodynamics has been introduced to describe how epidemiological, immunological and evolutionary processes shape pathogen phylogenies (14,15). Here, we hence introduced in essence a new phylodynamic framework describing how the size of polytomies is influenced by epidemic dynamics. Importantly, our approach could help to interpret sequence data in the early stage of an outbreak when genetic diversity is still limited (e.g. below the phylodynamic threshold) (44) and phylogenetic uncertainty is high. This may though require adapting our method to account for right-truncated clusters of identical sequences.

Detecting changes in the reproduction number and in the transmissibility of genetic variants is a crucial element of epidemic preparedness. Former modelling efforts have underlined the value of monitoring anomalous epidemiological cluster sizes for epidemic surveillance (9,43). Here, we developed an analogous framework to determine whether a genetic variant was characterized by a different reproduction number. This was done by comparing the size distribution of clusters of identical sequences between two pathogen genetic populations. Interestingly, a simple visual inspection of these distributions for the Alpha, Delta and Omicron variants of concerns (VOCs) in Washington state was already suggesting that VOCs were associated with larger polytomy sizes (Figure S12). This means that it should be possible to set up a surveillance system monitoring the size of clusters of identical sequences to detect anomalous chains, which may be attributable to changes in the reproduction number or increased transmissibility of genetic variants. Our framework could also easily be adapted to assess whether a variant is associated with a different dispersion parameter.

Our modelling framework relies on the assumption that two identical sequences are always linked within an epidemiological cluster. This hypothesis could be broken if an identical sequence was introduced multiple times in a given population. To account for such repeated introductions, our likelihood calculation could easily be extended to integrate over the potential sub-clusters of identical sequences by adapting the combinatorial approach proposed by Blumberg and Lloyd-Smith (9). We also assumed that no convergent evolution was occurring. Estimates could also be biased if the observed cluster size distribution was not representative of the true underlying cluster size distribution. For example, if larger clusters tended to be overrepresented in the analyzed dataset, *R* estimates could be biased upwards. This could result from case investigation favoring the detection of larger clusters or from sequencing biased towards individuals with lower cycle threshold (Ct) values, who might be more infectious.

Finally, our inference framework is based on some assumptions regarding the probability *p* that two members of a transmission pair have the same consensus sequence. Here, we estimated this probability by the one that a transmission event occurs before a substitution one. Instead of a simulation approach, sequencing data in transmission pairs can directly inform estimates of *p*. Using deep-sequencing data from SARS-CoV-2 and influenza household transmission pairs, we obtained estimates within a similar range than with our simulation approach (Figure S14). Our simulation approach would result in biased *p* estimates for pathogens with longer generation times and large transmission bottlenecks (Supplementary text A, Figures S17-S19). We also approximated the evolutionary time between two samples from a transmission pair by the generation time. In practice, the timing between transmission and sampling of both the donor and the recipient impacts the evolutionary time between samples. For specific case studies, information regarding the delay between infection and sequencing, when available, could be included to improve estimates of *p*. Additionally, we estimated *p* by relying on substitution rates drawn from the literature, which are typically obtained through phylogenetic analyses whose timescales span longer than that of transmission events. As viral evolutionary rates are impacted by their measurement timescales (with higher rates estimates over shorter timescales) (45), estimates over longer timescales drawn from the literature might underestimate the evolutionary rate at the transmission event scale, which could result in *p* estimates biased upwards. Relying on estimates of the evolutionary rate obtained at the transmission event timescale, if available, would be valuable. The comparison between our estimates and estimates from household transmission pairs however suggests that relying on estimates over longer timescales does not yield unreasonable results.

Our work opens up a number of exciting research directions, such as accounting for non-stationary epidemic processes (*e.g.* with reproduction numbers and sequencing fraction varying over time) (2), considering more complex observation processes (*e.g.* for pathogens where case investigation might increase the likelihood of observing larger clusters) (10), evaluating how heterogeneity in infectious duration is susceptible to impact transmission heterogeneity estimates (46), or extending our approach to pathogens responsible for longer infections characterized by considerable within-host diversity (*e.g.* by assessing how the reproduction number and the dispersion parameter may impact the pathogen genetic diversity at the population level (47)). We also showed that when the reproduction number reaches the threshold of 1/*p*, where *p* is the probability that an infector and an infectee have the same consensus sequence, our approach was no longer valid. Previous methods relying on the size of epidemiological clusters were valid when *R < 1* (9). Here, we have extended this threshold by looking at genomic clusters. Overall, future research extending our framework to values of *R* lying above the threshold (*e.g.* by analyzing the subset of clusters that got extinct (see Supplementary text B, Figures S20-S23), by also including the sequence collection date or by analyzing right truncated cluster size distributions) would be of primary interest.

Building on the observation that clusters of identical sequences are nested within epidemiological clusters, we introduced a novel statistical framework to (i) infer the reproduction number and transmission heterogeneity from pathogen genomes and (ii) determine whether a specific variant subpopulation is characterized by a transmission advantage. Our method is suitable to analyse epidemics even when establishing epidemiological relationships is difficult (*e.g.* vector-borne infections), which was the foundation of previous methods used to quantify transmission heterogeneity, hence constituting a valuable new tool to study current epidemics and prepare for future ones.

## Material and methods

### Offspring with identical genomes distribution

We used a branching process formulation to derive the distribution of the number of offspring with identical genomes. Let *Z* (respectively *Z̅*) be a random variable corresponding to the number of offspring generated by a single infected individual (respectively the number of offspring with identical genomes). Let *g* and *g̅* denote their respective probability generating function:

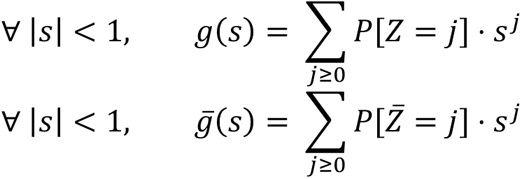

We introduced *p*, the probability that an infected individual has the same consensus genome as their infector. We approximated the evolutionary time between two samples from a transmission pair by the generation time. For pathogens causing acute infections characterized by narrow transmission bottlenecks, *p* can be approximated by the probability that a transmission event occurs before a substitution event (Supplementary text A). The distribution of the number of offspring with identical genomes *Z̅* can then be related to that of the number of offspring stemming from a single infector through a binomial distribution:

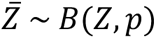

This enabled us to derive the following relationship between *g* and *g̅*:

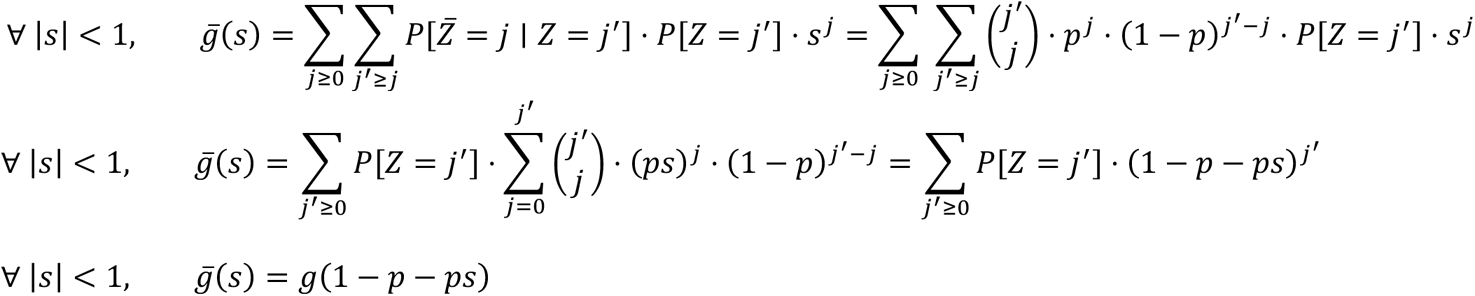

Following Lloyd-Smith et al.(5), we assumed that the offspring distribution (*Z*) follows a negative binomial distribution of mean *R* and dispersion parameter *k*. The probability generating function *g* of *Z* thus had the following form:

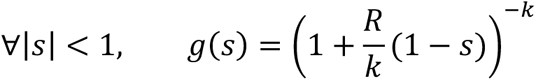

The probability generating function *g̅* of *Z̅* could then be derived as:

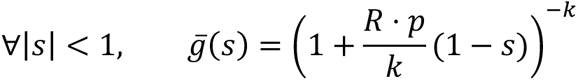

which is the probability generating function of a negative binomial distribution of mean *p* ⋅ *R* and dispersion parameter *k*. Hence:

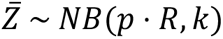

### Distribution of the size of clusters of identical sequences

Nishiura et al (8) and Blumberg & Lloyd-Smith (9) developed an inference framework to estimate the reproduction number and the dispersion parameter from the size of epidemiological clusters. Here, we extended this to estimate these two parameters from the size distribution of clusters of identical sequences. Let *r_j_* denote the probability for the size of a cluster of identical sequences to be equal to *j*. We have:

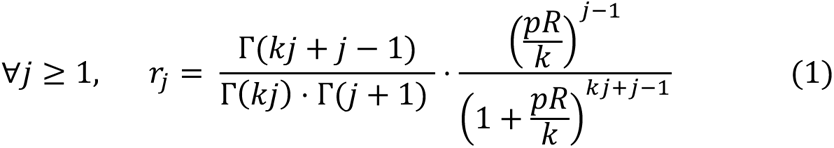

### Evaluating the contribution of highly infectious individuals to the size of clusters of identical sequences

We quantified the contribution of a cluster’s most infectious individual to the size of that cluster of identical sequences. We defined the most infectious individual as the cluster member that generated the most offspring. To do so, we simulated a branching process with substitution. At each generation, we drew the number of new individuals infected by a given individual from a negative binomial distribution of parameters (*R*, *k*). This enabled us to identify the most infectious individual in the cluster. We then drew whether the new infected individuals were infected by the same genotype that their infector from a Bernoulli distribution of parameter *p*.

### Accounting for the partial sequencing of infections

In practice, clusters will only be partially observed as (i) solely a fraction of infections may be detected by the surveillance system (we refer to the detected infections as cases) and (ii) solely a fraction of cases will then be sequenced. This means that the observed distribution of the size of clusters of identical sequences will differ from the true underlying size distribution of clusters of identical sequences. Failing to account for this partial observation has been shown to lead to biased estimates of *R* and *k* when inferring them from the size distribution of epidemiological clusters (10). Here, we explicitly modeled the cluster observation process.

Let *ρ* denote the probability that a given infected individual is sequenced. Let *s_j_* denote the probability to observe *j* identical sequences among an arbitrary cluster of identical sequences. We have:

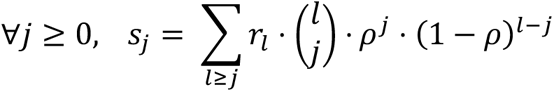

As clusters of size 0 are never observed, we were interested in the probability 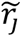 for an observed cluster to be of a given size *j* knowing in was observed (which corresponds to the probability for an observed cluster to be of size *j* conditional on this cluster being of size greater or equal to 1):

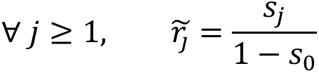

In practice, we approximated *s*_*j*_ with a truncated sum, assuming cluster sizes remained below a certain threshold *c_max_*:

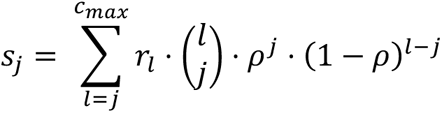

### Statistical framework

We used a likelihood-based approach to estimate *R* and *k* from the size distribution of clusters of identical sequences. Let *D* denote data describing the size of clusters of identical sequences. Let *N* denote the number of distinct clusters in *D* and *n*_*j*_ be the number of clusters of size *j*. The log-likelihood of the data was derived as:

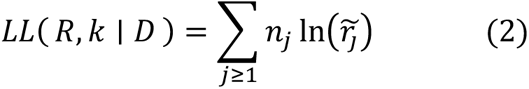

Maximum likelihood estimates were obtained using a constrained quasi-Newton optimization approach imposing values of the reproduction number ranging between 0.01 and 10.0 and values of the dispersion parameter ranging between 0.001 and 10.0. This was done using the *optim* base R function (48).

Confidence intervals were obtained by likelihood profiling (9). Let *LL^MLE^* denote the maximum log-likelihood of the log-likelihood function of (*R*, *k*) defined in (2). We introduced the profile log-likelihood of *R* as follows:

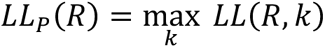

A confidence interval associated with a confidence level of 1 − *α* then corresponds to values of *R* verifying:

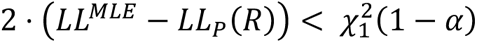

where 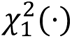 is the quantile function of a chi-squared distribution with 1 degree of freedom. Confidence intervals for the dispersion parameter *k* were obtained in a similar manner, by inverting the role of *R* and *k* above. The bounds of the confidence intervals were considered unresolved when lying outside the range of 0.01-10 for *R* and 0.001-10 for *k*.

### Simulation approach to estimate the probability that transmission occurs before substitution using a more flexible framework for the generation time

We used a simulation approach to estimate the probability that a transmission event occurs before a substitution one. More details on the simulation approach are available in the Supplement. Parameters used in the simulations along the resulting probability estimate are detailed in Tables S1-S2. A visual comparison of the distribution of the delay before a substitution and a transmission event is available in Figure S15.

### Using transmission pair data to estimate the proportion of transmission pairs with identical consensus sequences

We compared the estimates we obtained for the probability p from our simulation approach with those stemming from the analysis of household transmission pair data (for both seasonal influenza (51) and SARS-CoV-2 (52)). From the data publicly available associated with the work of McCrone et al. (51), we identified 38 H3N2 transmission pairs (23 had identical consensus sequences). Re-analyzing the data of Bendall et al. (52), we identified 79 pre-Omicron transmission pairs (47 had identical consensus sequences) and 53 transmission pairs infected by the Omicron variant (47 had identical consensus sequences).

### Simulation study

To evaluate the performance of our novel method, we applied our inference framework to synthetic clusters of identical sequences. Each simulation scenario was characterized by:

- the probability *p* that an infector and an infectee have the same consensus genome (38%, 50% and 83%),
- the proportion of infections sequenced *ρ* (50%, 10% or 1%),
- the reproduction number *R* (ranging between 0.5 and 3.0),
- the dispersion parameter *k* (ranging between 0.01 and 1.0).

Clusters of identical sequences were generated using a branching process in which the number of offspring with identical sequences was drawn from a negative binomial distribution of mean *R* ⋅ *p* and dispersion parameter *k*. As some clusters may never get extinct, clusters were simulated until reaching a size of 10,000. We then accounted for the partial sequencing of infections by drawing the observed cluster sizes from a binomial distribution with probability parameter *ρ*.

For each parameter combination, we generated datasets comprised of 50, 100, 1000 or 5000 clusters of identical sequences. For each of these datasets, we then jointly inferred the value of the reproduction number and the dispersion parameter *k* using our inference framework. The estimated values were then compared with the ones used to generate the clusters.

### Inference framework to quantify the transmission advantage of genetic variants

We next extended our inference framework to study the transmission advantage of a specific genetic variant. This was done by performing a likelihood ratio test to determine whether the size distribution of clusters of identical pathogen sequences for a specific variant (superscript *V*) corresponds to a different reproduction number than the size distribution of clusters of identical pathogen sequences for non-variant sequences (superscript *NV*). Similar methods have been used to monitor changes in the reproduction number from epidemiological cluster data (9). We assumed that the variant and the non-variant pathogen had the same dispersion parameter *k*.

More specifically, let *D^V^* (respectively *D^NV^*) denote the data describing the size of cluster of identical sequences for the variant (respectively the non-variant). We first computed the log-likelihood of the combined dataset *D_combined_* = (*D^V^*, *D^NV^*) assuming that the variant and the non-variant were both characterized by the reproduction number *R* and the dispersion parameter *k*: *LL_combined_*(*R*, *k* |*D_combined_*). We then estimated the maximum-likelihood estimates of this combined likelihood 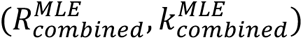. Second, we computed a splited log-likelihood, this time assuming that the variant and the non-variant were respectively characterized by the reproduction numbers *R^V^* and *R^NV^*:

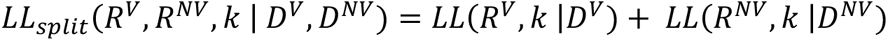

We then estimated the corresponding maximum likelihood estimates 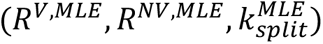. As the combined model is nested within the split one, we performed a likelihood ratio test by computing the following test statistic:

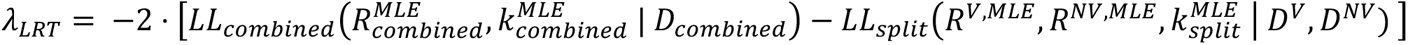

We then derived the corresponding p-value under a chi-squared distribution with 1 degree of freedom. In all the analyses reported in the manuscript, we used a type I error (Alpha risk) of 5%.

### Simulation study to evaluate the performance of our inference framework

We used a simulation study to ascertain the performance of our inference framework. Synthetic cluster data were generated from a branching process and exploring a range of assumptions regarding:

- the variant transmission advantage (defined as 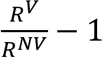)
- the probability that an infector and an infectee have the same consensus sequence
- the dispersion parameter
- the reproduction number of the non-variant
- the number of clusters simulated for both the variant and the non-variant (50, 100, 1000, 5000)

For each combination of parameters, we evaluated the sensitivity of our statistical framework in detecting a difference between the reproduction numbers of two genetic subpopulations. This was done running our inference framework on 100 different datasets generated using the same set of parameters. For each combination of parameters, we then computed the sensitivity as the fraction of simulations for which we were able to detect a transmission advantage using a significance level of 5%. We also computed the absolute transmission advantage bias as the difference between the estimated transmission advantage (maximum-likelihood estimate) and the true one.

### Application to different epidemiological situations

We applied our novel frameworks to the following datasets:

- Sequences from the 2013-2015 Middle East respiratory syndrome outbreak (26)
- Sequences from the 2017-2018 measles outbreak in the Veneto region (Italy) (30)
- Sequences from the COVID-19 pandemic in New Zealand during the Zero COVID era (April 2020- July 2021) obtained from the GISAID EpiCoV database (53,54).
- SARS-CoV-2 sequences from Washington State obtained from the GISAID EpiCoV database (53,54). We applied our transmission advantage framework to this dataset.

A detailed description of these datasets and of the different scenarios explored is available in the Supplementary Information (section Supplementary methods – Application to different epidemiological situations).

### Defining clusters of identical sequences

For each dataset, we computed a pairwise distance matrix between aligned sequences using the *ape* R package (55). If there weren’t any missing data (sites) in the sequences, this matrix would directly enable to reconstruct clusters of identical sequences. In practice, this is not the case. We thus defined clusters of identical sequences as maximal groups of sequences for which all sequences were at a null distance to all other sequences within the cluster. The difference between pairwise distances and clusters of identical sequences is illustrated in Figure S16 for MERS-CoV. The cluster generation was done using the R *igraph* package (56).

Due to the large number of sequences available for SARS-CoV-2, generating a single pairwise genetic distance matrix would be computationally and memory intensive. Instead, we grouped sequences by Pango lineage assigned with Nextclade (34,35) and generated a pairwise genetic distance matrix for each Pango lineage.

## Supporting information

Acknowledgments table for SARS-CoV-2 New Zealand sequences

Acknowledgments table for SARS-CoV-2 Washington state sequences

## Data Availability

The codes and data used in this paper can be found at https://github.com/blab/size-genetic-clusters.

## Data and code accessibility

Code and data used in this paper can be found at https://github.com/blab/size-genetic-clusters. The MERS-CoV aligned sequences used in the analysis were directly extracted from the aligned sequence data available at (57) for human infections. The GISAID accession numbers associated with the SARS-CoV-2 sequences used in this analysis (both for New-Zealand and Washington state) are provided at https://github.com/blab/size-genetic-clusters/tree/main/data. The size distributions of clusters of identical sequences used to run the different analyses are available in the associated GitHub repository. We provide scripts to ingest FASTA files to produce cluster distributions and scripts to estimate R and k from an input cluster distribution.

## Author contributions

CTK and TB conceived the study. CTK developed the methods, conducted the analyses, interpreted the results, and wrote the first draft. CTK and TB edited the initial manuscript.

## Acknowledgments

We would like to thank James Hadfield for his help on the interpretation of the New Zealand data, John Huddleston, Jennifer Chang, Alli Black and Paolo Bosetti for helpful feedback on a first draft and Simon Cauchemez for helpful discussions. We gratefully acknowledge all data contributors, i.e., the Authors and their Originating laboratories responsible for obtaining the specimens, and their Submitting laboratories for generating the genetic sequence and metadata and sharing via the GISAID Initiative, on which this research is based. We also greatly acknowledge authors, alongside their originating and submitting laboratories for generating and submitting the sequences and associated metadata on GenBank. The GenBank accession numbers associated with the measles sequences used in the analysis are detailed in Table S6. Acknowledgement tables for SARS-CoV-2 sequences are available at https://github.com/blab/size-genetic-clusters/tree/main/data.

