## Supplementary information for "Estimating the reproduction number and transmission heterogeneity from the size distribution of clusters of identical pathogen sequences"

**Supplementary methods – Application to different epidemiological situations**

**Supplementary methods – Estimating the probability that a transmission event occurs before a substitution event**

**Supplementary tables S1-S9**

**Supplementary figures S1-S23**

**Supplementary text**

**A - Impact of infectious duration and transmission bottleneck size on the proportion of transmission pairs with identical consensus sequences**

**B - Inference of transmission parameters conditional on cluster extinction**

**References**

**Supplementary methods – Application to different epidemiological situations**

**2013-2015 Middle East respiratory syndrome outbreak**

Between March 2012 and 31 December 2015, 1,646 cases were identified globally (49). We analyzed 174 aligned MERS-CoV sequence data sampled in humans between 2013 and 2016 analyzed in Dudas et al (25) and made available at (50). This set of sequences thus corresponded to 10.6% of all detected cases. For the analysis, we explored scenarios where:

(i) all infections were detected(corresponding to an infection sequencing ratio of 10.6%)
(ii) half of infections were detected (corresponding to an infection sequencing ratio of 5.3%)

For the MERS analysis, we used a threshold $c_{max}$ for the size of cluster of 10,000. Increasing this value to 50,000 did not impact our estimates.

**2017-2018 measles outbreak in the Veneto Region (Italy)**

We used data describing the 2017-2018 measles outbreak in the Veneto Region, located in the North-East of Italy (4). We used 30 sequences (out of 322 suspected cases) analyzed in Pacenti et al (4). Sequences were aligned using the Nextstrain measles workflow (5–7). This set of sequences thus corresponded to 9.3% of detected cases. For the analysis, we explored scenarios where:

(i) all infections were detected (corresponding to an infection sequencing ratio of 9.3%)
(ii) half of infections were detected (corresponding to an infection sequencing ratio of 4.7%)

For the measles analysis, we used a threshold $c_{max}$ for the size of cluster of 10,000. Increasing this value to 50,000 did not impact our estimates.

**COVID-19 pandemic in New-Zealand**

We analyzed 27,565 SARS-CoV-2 sequences from New Zealand downloaded from the GISAID EpiCoV database on December 8$^{th}$, 2022 (8, 9) and curated using the Nextstrain nCoV ingest pipeline (10). Clusters of identical sequences were generated using the approach detailed below and grouped by time period based on the collection date of the earliest sequence within the cluster.

We estimated the offspring distribution of COVID-19 in New Zealand during the Zero COVID era (April 2020 – July 2021). We assumed that the dispersion parameter *k* remained constant throughout the period but allowed the reproduction number *R* to vary between time periods (April-May 2020, June-December 2020, January-April 2021 and May-July 2021). As a baseline scenario, we considered that throughout this period, 80% of infections were detected. Since autochthonous transmission remained extremely limited during this period, it is indeed unlikely that a large fraction of infections was undetected by the surveillance system. As a sensitivity analysis, we also explored scenarios where 50% and 100% of infections were detected. For each time period, the fraction of cases sequenced was estimated as the ratio of the number of sequences collected and of the number of cases (11) reported during this time period.

For this analysis, we used a threshold $c_{max}$.for the size of cluster of 10,000. Increasing this value to 50,000 did not impact our estimates.

As k estimates might be difficult to interpret, we computed the expected proportion of individuals contributing to 80% of transmission events. This was done for maximum-likelihood estimates of the dispersion parameter and the reproduction numbers across the 4 periods (central estimates) and for the bounds of 95% likelihood profile confidence interval for $k$ (while using maximum likelihood estimates for R).

**COVID-19 pandemic in Washington state**

We analyzed 140,790 SARS-CoV-2 sequences from Washington state (United States of America) downloaded from the GISAID EpiCoV database on December 8th, 2022 (8, 9) and curated using the Nextstrain nCoV ingest pipeline (10). As for the New-Zealand analysis, we allocated a date to each cluster of identical sequences based on the collection date of the earliest sequence within that cluster.

We applied our transmission advantage framework to the following variants: D614G, Epsilon, Alpha, Delta, Omicron BA.1, Omicron BA.2 and Omicron BA.4/BA.5. We defined variant-specific study periods beginning on the day when at least 10 variant sequences have been collected (cumulative) and lasting between 1 and 60 days (Table S8). For the Omicron BA.1 analysis, we only considered time-windows of 50 days maximum, to restrict analyses before the spread of BA.2. For each window of analysis, we selected the clusters of identical SARS-CoV-2 sequences who started during this period. We generated the size distribution of clusters of identical sequences for both the variant and non-variant genetic sub-populations (Figure S12) and only considered clusters who were initiated during this time window. From this, we applied tour transmission advantage inference framework and computed a p-value associated with the statistical test defined by the following null hypothesis:

$H_0$: There is no difference in the reproduction number of the variant and non-variant.

This was done by accounting for the fraction of cases sequenced in Washington state during these different time periods and exploring different assumptions regarding the fraction of infections detected. We also accounted for the shorter generation time associated with Omicron variants (12, 13), which translated on the probability that an infector and an infectee have the same consensus sequence (Table S2).

**Supplementary methods – Estimating the probability that a transmission event occurs before a substitution event**

The generation time can often not be approximated using an exponential distribution (e.g. its variance is generally not equal to its mean). We relaxed this assumption and empirically derived the probability that a transmission event occurs before a substitution one using a simulation approach for the following pathogens: mumps virus, MERS-CoV, SARS-CoV, Ebola virus, mpox during the 2022-2023 outbreak, measles virus, RSV, Zika virus, influenza A (H1N1pdm and H3N2) and SARS-CoV-2. For SARS-CoV-2, we accounted for a reduction in the generation time for Omicron variant (49,50). For each pathogen, we identified relevant parameters describing the mean and the standard deviation of the generation time of these pathogens. The generation time describes the average duration between the infection time of an index case and the time at which this primary case infects a secondary case. We then drew $n_{sim} = 10^7$ generation intervals from a Gamma distribution parametrized with the same mean and standard deviation. We also drew $n_{sim} = 10^7$ delays until the occurrence of a first substitution for these different pathogens from an exponential distribution of rate the substitution rate obtained from the literature. We then computed the proportion of simulations for which the generation time was shorter than the delay until the occurrence of a first substitution to obtain an estimate of the probability that transmission occurs before substitution.

We explored how uncertainty around the mean generation time and the pathogen's substitution rate impacted estimates of p. Uncertainty in the substitution rate (respectively the mean generation time) was accounted for by fixing the mean generation time (respectively the substitution rate) to the central estimate and using the lower and upper bound for the substitution rate (respectively the generation time) reported in the different studies. From this, we obtained 4 values for the probability that transmission occurs before substitution. We reported the lowest and the highest of these 4 estimates as our lower and upper bound estimates for *p*.

**Table S1: Estimates of the probability that transmission occurs before subtitution for different pathogens along assumptions for the generation time distribution and the substitution rate used for the estimation.** The numbers in parentheses correspond to uncertainty ranges.

| Pathogen | Generation time in days | | Substitution rate in subs/site/ year (uncertainty range) | Genome length | Ref. for the generation time | Ref. for the substitution rate |
|---|---|---|---|---|---|---|
| | Mean (uncertainty range) | Standard deviation (days) | | | | |
| MERS-CoV | 6.8 (6.0-7.8) | 6.3 | $4.81\cdot10^{-4}$ ($2.74\cdot10^{-4}$-$6.88\cdot10^{-4}$) [A] | 30130 | (14) | (15) |
| Measles virus | 11.7 (9.9-13.8) [B] | 1.8 | $5.13\cdot10^{-4}$ ($4.84\cdot10^{-4}$-$5.13\cdot10^{-4}$) [C] | 15894 | (16) | (17) |
| Ebola virus | 14.2 (13.1-15.5) | 7.1 | $9.82\cdot10^{-4}$ ($9.01\cdot10^{-4}$–$10.6\cdot10^{-4}$) | 18958 | (18) | (19) |
| Zika virus | 20.0 (15.6-25.6) | 7.4 | $1.12\cdot10^{-3}$ ($0.97\cdot10^{-3}$-$1.27\cdot10^{-3}$) | 10274 | (20) | (21) |
| Mpox virus (2022-2023 outbreak) | 12.5 (7.5-17.3) | 5.7 | $8.41\cdot10^{-5}$ ($7.71\cdot10^{-5}$ -$9.10\cdot10^{-5}$) | 197209 | (22) | (23) |
| Influenza A (H1N1) | 2.6 (2.2-3.5) | 1.3 | $3.41\cdot10^{-3}$ ($3.15\cdot10^{-3}$-$3.67\cdot10^{-3}$) | 13154 | (24) | (25) |
| Influenza A (H3N2) | 2.8 (3.1-4.6) | 2.0 | $1.43\cdot10^{-3}$ ($1.41\cdot10^{-3}$-$1.44\cdot10^{-3}$) | 13486 | (26) | Obtained from the slope of a linear regression of number of mutations accumulated vs time |
| Mumps virus | 18.0 (17.4-18.6) | 3.5 | $8.6\cdot10^{-4}$ ($5.06\cdot10^{-4}$-$12.7\cdot10^{-4}$) | 15384 | (16) | (19) |
| RSV-A | 7.5 (7.0-8.1) | 2.1 | $6.47\cdot10^{-4}$ ($5.56\cdot10^{-4}$ - $7.38\cdot10^{-4}$) | 15200 | (16) | (27) |
| SARS-CoV | 8.7 [D] | 3.6 | $2.08\cdot10^{-3}$ ($0.8\cdot10^{-3}$-$2.38\cdot10^{-3}$) | 29714 | (28) | (29) |
| SARS-CoV-2 (pre-Omicron) | 5.9 (5.2-7.0) | 4.8 | $1.10\cdot10^{-3}$ ($7.03\cdot10^{-4}$ -$1.50\cdot10^{-3}$) | 29500 | (30) | (31) |
| SARS-CoV-2 (Omicron) | 4.9 (4.2-6.0) [E] | 4.8 | | | (12, 13) | |

[A] The uncertainty range for the MERS-CoV substitution rate was obtained by subtracting and adding the standard deviation reported in (16) to the central estimate.

[B] The uncertainty range for the measles generation time was obtained by considering the range of values reported for the mean measles generation time in (17).

[C] The uncertainty range for the measles' substitution rate was obtained by subtracting and adding the standard deviation obtained with the Nextstrain measles workflow (32).

[D] We did not explore any uncertainty around the SARS mean generation time (no estimates found).

[E] For Omicron, we assumed that the mean generation time was one day shorter than for pre-Omicron variants (12, 13) and considered that it was characterized by the same standard deviation.

**Table S2: Estimates of the probability that transmission occurs before substitution for different pathogens.**

| Pathogen | Values used to inform the generation time / the substitution rate | | | | | Central estimate (uncertainty range) |
|---|---|---|---|---|---|---|
| | Central / Central | Lower/ Central | Upper / Central | Central / Lower | Central / Upper | |
| MERS-CoV | 0.78 | 0.81 | 0.75 | **0.87** | **0.72** | 0.78 (0.72-0.87) |
| Measles virus | 0.77 | **0.80** | **0.74** | 0.78 | 0.77 | 0.77 (0.74-0.80) |
| Ebola virus | 0.51 | **0.54** | **0.48** | 0.54 | 0.49 | 0.51 (0.48-0.54) |
| Zika virus | 0.55 | **0.63** | **0.46** | 0.59 | 0.51 | 0.55 (0.46-0.63) |
| Mpox virus (2022-2023 outbreak) | 0.59 | **0.73** | **0.47** | 0.61 | 0.56 | 0.59 (0.47-0.73) |
| Influenza A (H1N1) | 0.74 | **0.77** | **0.66** | 0.75 | 0.72 | 0.74 (0.66-0.77) |
| Influenza A (H3N2) | 0.82 | **0.85** | **0.79** | 0.83 | 0.82 | 0.82 (0.79-0.85) |
| Mumps virus | 0.53 | 0.54 | 0.51 | **0.68** | **0.39** | 0.53 (0.39-0.68) |
| RSV-A | 0.82 | 0.83 | 0.81 | **0.84** | **0.80** | 0.82 (0.80-0.84) |
| SARS-CoV | 0.27 | - | - | **0.58** | **0.23** | 0.27 (0.23-0.58) |
| SARS-CoV-2 (pre-Omicron) | 0.64 | 0.68 | 0.58 | **0.74** | **0.56** | 0.64 (0.56-0.74) |
| SARS-CoV-2 (Omicron) | 0.69 | 0.74 | 0.63 | **0.78** | **0.63** | 0.69 (0.63-0.78) |

**Table S3: Parameter estimates for MERS.** Maximum likelihood estimates are reported along 50% and 95% confidence intervals (CI).

| Estimate used for the probability that transmission occurs before substitution | Proportion of infections detected as cases | Reproduction number *R* estimate | Dispersion parameter *k* estimate |
|---|---|---|---|
| Central | 1.0 | 0.57<br>50%CI: (0.54-0.61)<br>95%CI: (0.46-0.70) | 0.14<br>50%CI: (0.09-0.20)<br>95%CI: (0.04-0.46) |
| | 0.5 | 0.65<br>50%CI: (0.61-0.68)<br>95%CI: (0.54-0.77) | 0.09<br>50%CI: (0.07-0.13)<br>95%CI: (0.03-0.26) |
| Lower bound from uncertainty range | 1.0 | 0.63<br>50%CI: (0.59-0.67)<br>95%CI: (0.50-0.77) | 0.14<br>50%CI: (0.09-0.20)<br>95%CI: (0.04-0.46) |
| | 0.5 | 0.71<br>50%CI: (0.67-0.75)<br>95%CI: (0.59-0.84) | 0.09<br>50%CI: (0.07-0.13)<br>95%CI: (0.03-0.26) |
| Upper bound from uncertainty range | 1.0 | 0.52<br>50%CI: (0.46-0.56)<br>95%CI: (0.42-0.64) | 0.14<br>50%CI: (0.09-0.20)<br>95%CI: (0.04-0.46) |
| | 0.5 | 0.59<br>50%CI: (0.55-0.62)<br>95%CI: (0.49-0.70) | 0.09<br>50%CI: (0.07-0.13)<br>95%CI: (0.03-0.26) |

**Table S4: Parameter estimates for measles.** Maximum likelihood estimates are reported along 50% and 95% confidence intervals (CI).

| Estimate used for the probability that transmission occurs before substitution | Proportion of infections detected as cases | Reproduction number *R* estimate | Dispersion parameter *k* estimate |
|---|---|---|---|
| Central | 1.0 | 0.58<br>50%CI: (0.47-0.73)<br>95%CI: (0.29-1.18) | 0.04<br>50%CI: (0.016-0.092)<br>95%CI: (0.003-0.45) |
| | 0.5 | 0.62<br>50%CI: (0.50-0.76)<br>95%CI: (0.32-1.17) | 0.02<br>50%CI: (0.009-0.05)<br>95%CI: (0.002-0.19) |
| Lower bound from uncertainty range | 1.0 | 0.61<br>50%CI: (0.49-0.77)<br>95%CI: (0.31-1.23) | 0.04<br>50%CI: (0.016-0.092)<br>95%CI: (0.003-0.45) |
| | 0.5 | 0.65<br>50%CI: (0.52-0.80)<br>95%CI: (0.34-1.23) | 0.02<br>50%CI: (0.009-0.05)<br>95%CI: (0.002-0.19) |
| Upper bound from uncertainty range | 1.0 | 0.56<br>50%CI: (0.45-0.71)<br>95%CI: (0.28-1.13) | 0.04<br>50%CI: (0.016-0.092)<br>95%CI: (0.003-0.45) |
| | 0.5 | 0.59<br>50%CI: (0.31-1.13)<br>95%CI: (0.48-0.73) | 0.02<br>50%CI: (0.009-0.05)<br>95%CI: (0.002-0.19) |

**Table S5: Parameter estimates for SARS-CoV-2 in New Zealand under our central estimate for the probability *p* that transmission occurs before substitution.** Maximum likelihood estimates are reported along 50% and 95% confidence intervals (CI).

| Proportion of infections detected as cases | Period | Reproduction number *R* estimate | Dispersion parameter *k* estimate |
|---|---|---|---|
| 1.0 | April – May 2020 | 0.82<br>95%CI: (0.65-1.01)<br>50%CI: (0.76-0.88) | 0.63<br>95%CI: (0.34-1.56)<br>50%CI: (0.5-0.82) |
| | June – December 2020 | 0.87<br>95%CI: (0.75-1.01)<br>50%CI: (0.83-0.91) | |
| | January – April 2021 | 0.74<br>95%CI: (0.61-0.90)<br>50%CI: (0.70-0.79) | |
| | May – July 2021 | 0.66<br>95%CI: (0.48-0.87)<br>50%CI: (0.60-0.72) | |
| 0.8 | April – May 2020 | 0.87<br>95%CI: (0.70-1.06)<br>50%CI: (0.82-0.93) | 0.62<br>95%CI: (0.33-1.54)<br>50%CI: (0.49-0.81) |
| | June – December 2020 | 0.92<br>95%CI: (0.80-1.05)<br>50%CI: (0.88-0.96) | |
| | January – April 2021 | 0.80<br>95%CI: (0.66-0.95)<br>50%CI: (0.75-0.84) | |
| | May – July 2021 | 0.71<br>95%CI: (0.54-0.92)<br>50%CI: (0.65-0.78) | |
| 0.5 | April – May 2020 | 0.98<br>95%CI: (0.82-1.14)<br>50%CI: (0.92-1.03) | 0.59<br>95%CI: (0.31-1.43)<br>50%CI: (0.47-0.77) |
| | June – December 2020 | 1.02<br>95%CI: (0.91-1.13)<br>50%CI: (0.98-1.05) | |
| | January – April 2021 | 0.90<br>95%CI: (0.78-1.04)<br>50%CI: (0.86-0.94) | |
| | May – July 2021 | 0.82<br>95%CI: (0.65-1.02)<br>50%CI: (0.77-0.88) | |

**Table S6: Parameter estimates for SARS-CoV-2 in New Zealand under our <u>lower bound</u> estimate for the probability *p* that transmission occurs before substitution.** Maximum likelihood estimates are reported along 50% and 95% confidence intervals (CI).

| Proportion of infections detected as cases | Period | Reproduction number *R* estimate | Dispersion parameter *k* estimate |
|---|---|---|---|
| 1.0 | April – May 2020 | 0.94<br>95%CI: (0.74-1.16)<br>50%CI: (0.87-1.01) | 0.63<br>95%CI: (0.34-1.53)<br>50%CI: (0.50-0.82) |
| | June – December 2020 | 1.00<br>95%CI: (0.86-1.15)<br>50%CI: (0.95-1.05) | |
| | January – April 2021 | 0.85<br>95%CI: (0.69-1.03)<br>50%CI: (0.79-0.90) | |
| | May – July 2021 | 0.75<br>95%CI: (0.55-1.00)<br>50%CI: (0.68-0.83) | |
| 0.8 | April – May 2020 | 1.00<br>95%CI: (0.80-1.21)<br>50%CI: (0.93-1.07) | 0.62<br>95%CI: (0.32-1.51)<br>50%CI: (0.49-0.80) |
| | June – December 2020 | 1.05<br>95%CI: (0.92-1.20)<br>50%CI: (1.01-1.10) | |
| | January – April 2021 | 0.91<br>95%CI: (0.75-1.08)<br>50%CI: (0.86-0.96) | |
| | May – July 2021 | 0.81<br>95%CI: (0.61-1.05)<br>50%CI: (0.74-0.89) | |
| 0.5 | April – May 2020 | 1.11<br>95%CI: (0.93-1.30)<br>50%CI: (1.06-1.18) | 0.58<br>95%CI: (0.30-1.39)<br>50%CI: (0.46-0.76) |
| | June – December 2020 | 1.16<br>95%CI: (1.04-1.30)<br>50%CI: (1.12-1.21) | |
| | January – April 2021 | 1.03<br>95%CI: (0.88-1.19)<br>50%CI: (0.98-1.08) | |
| | May – July 2021 | 0.94<br>95%CI: (0.74-1.16)<br>50%CI: (0.87-1.01) | |

**Table S7: Parameter estimates for SARS-CoV-2 in New Zealand under our upper bound estimate for the probability *p* that transmission occurs before substitution.** Maximum likelihood estimates are reported along 50% and 95% confidence intervals (CI).

| Proportion of infections detected as cases | Period | Reproduction number *R* estimate | Dispersion parameter *k* estimate |
|---|---|---|---|
| 1.0 | April – May 2020 | 0.71<br>95%CI: (0.56-0.87)<br>50%CI: (0.66-0.76) | 0.64<br>95%CI: (0.34-1.58)<br>50%CI: (0.51-0.83) |
| | June – December 2020 | 0.75<br>95%CI: (0.65-0.87)<br>50%CI: (0.72-0.79) | |
| | January – April 2021 | 0.64<br>95%CI: (0.53-0.78)<br>50%CI: (0.60-0.68) | |
| | May – July 2021 | 0.57<br>95%CI: (0.42-0.75)<br>50%CI: (0.52-0.62) | |
| 0.8 | April – May 2020 | 0.75<br>95%CI: (0.61-0.91)<br>50%CI: (0.71-0.80) | 0.63<br>95%CI: (0.33-1.57)<br>50%CI: (0.50-0.82) |
| | June – December 2020 | 0.80<br>95%CI: (0.70-0.91)<br>50%CI: (0.76-0.83) | |
| | January – April 2021 | 0.69<br>95%CI: (0.57-0.82)<br>50%CI: (0.65-0.73) | |
| | May – July 2021 | 0.62<br>95%CI: (0.46-0.80)<br>50%CI: (0.56-0.67) | |
| 0.5 | April – May 2020 | 0.84<br>95%CI: (0.71-0.98)<br>50%CI: (0.80-0.89) | 0.60<br>95%CI: (0.31-1.47)<br>50%CI: (0.47-0.79) |
| | June – December 2020 | 0.88<br>95%CI: (0.79-0.98)<br>50%CI: (0.85-0.91) | |
| | January – April 2021 | 0.78<br>95%CI: (0.67-0.90)<br>50%CI: (0.75-0.82) | |
| | May – July 2021 | 0.71<br>95%CI: (0.56-0.88)<br>50%CI: (0.66-0.76) | |

**Table S8: Definitions of the study periods for the Washington state SARS-CoV-2 analysis.** Dates are reported in a YYYY-MM-DD format.

| Variant under study | Date of first collection of the variant | Date from which at least 10 variant sequences were collected (cumulative) | Corresponding Nextstrain clades |
|---|---|---|---|
| D614G | 2020-02-22 | 2020-03-10 | 19A, 19B, 19C |
| Epsilon | 2020-11-17 | 2020-12-13 | 21C (Epsilon) |
| Alpha | 2020-11-23 | 2021-01-18 | 20I (Alpha, V1) |
| Delta | 2021-04-03 | 2021-04-12 | 21A (Delta), 21I (Delta), 21J (Delta) |
| Omicron (BA.1) | 2021-09-29 | 2021-12-01 | 21K (Omicron) |
| Omicron (BA.2) | 2022-01-03 | 2022-01-12 | 21L (Omicron) |
| Omicron (BA.4, BA.5) | 2022-04-15 | 2022-05-08 | 22A (Omicron), 22B (Omicron) |

**Table S9: Genbank accession numbers for measles sequences used in the analysis.** All sequences were obtained from Pacenti et al. using the Nextstrain measles workflow (4, 7).

| Strain name | Accession number | URL |
|---|---|---|
| Padova.ITA/13.17/1/D8 | MK513623 | https://www.ncbi.nlm.nih.gov/nuccore/MK5136223 |
| Padova.ITA/14.17/3/D8 | MK513625 | https://www.ncbi.nlm.nih.gov/nuccore/MK513625 |
| Padova.ITA/16.17/4/B3 | MK513613 | https://www.ncbi.nlm.nih.gov/nuccore/MK513613 |
| Padova.ITA/14.17/7/B3 | MK513607 | https://www.ncbi.nlm.nih.gov/nuccore/MK513607 |
| Padova.ITA/16.17/2/B3 | MK513611 | https://www.ncbi.nlm.nih.gov/nuccore/MK513611 |
| Padova.ITA/16.17/3/B3 | MK513612 | https://www.ncbi.nlm.nih.gov/nuccore/MK513612 |
| Padova.ITA/14.17/2/D8 | MK513624 | https://www.ncbi.nlm.nih.gov/nuccore/MK513624 |
| Padova.ITA/16.17/6/B3 | MK513615 | https://www.ncbi.nlm.nih.gov/nuccore/MK513615 |
| Padova.ITA/20.17/1/B3 | MK513619 | https://www.ncbi.nlm.nih.gov/nuccore/MK513619 |
| Padova.ITA/14.17/4/B3 | MK513605 | https://www.ncbi.nlm.nih.gov/nuccore/MK513605 |
| Padova.ITA/13.17/1/B3 | MK513600 | https://www.ncbi.nlm.nih.gov/nuccore/MK513600 |
| Padova.ITA/21.17/1/B3 | MK513620 | https://www.ncbi.nlm.nih.gov/nuccore/MK513620 |
| Padova.ITA/19.17/1/B3 | MK513617 | https://www.ncbi.nlm.nih.gov/nuccore/MK513617 |
| Padova.ITA/14.17/2/B3 | MK513603 | https://www.ncbi.nlm.nih.gov/nuccore/MK513603 |
| Padova.ITA/14.17/5/B3 | MK513606 | https://www.ncbi.nlm.nih.gov/nuccore/MK513606 |
| Padova.ITA/16.17/1/B3 | MK513610 | https://www.ncbi.nlm.nih.gov/nuccore/MK513610 |
| Padova.ITA/13.17/2/B3 | MK513601 | https://www.ncbi.nlm.nih.gov/nuccore/MK513601 |
| Venezia.ITA/22.17/3/D8 | MK513627 | https://www.ncbi.nlm.nih.gov/nuccore/MK513627 |
| Padova.ITA/19.17/2/B3 | MK513618 | https://www.ncbi.nlm.nih.gov/nuccore/MK513618 |
| Padova.ITA/11.17/1/B3 | MK513598 | https://www.ncbi.nlm.nih.gov/nuccore/MK513598 |
| Padova.ITA/24.17/1/B3 | MK513622 | https://www.ncbi.nlm.nih.gov/nuccore/MK513622 |
| Padova.ITA/15.17/1/B3 | MK513608 | https://www.ncbi.nlm.nih.gov/nuccore/MK513608 |
| Padova.ITA/21.17/2/B3 | MK513621 | https://www.ncbi.nlm.nih.gov/nuccore/MK513621 |
| Padova.ITA/14.17/1/B3 | MK513602 | https://www.ncbi.nlm.nih.gov/nuccore/MK513602 |
| Padova.ITA/15.17/2/B3 | MK513609 | https://www.ncbi.nlm.nih.gov/nuccore/MK513609 |
| Padova.ITA/14.17/3/B3 | MK513604 | https://www.ncbi.nlm.nih.gov/nuccore/MK513604 |
| Padova.ITA/17.17/3/B3 | MK513616 | https://www.ncbi.nlm.nih.gov/nuccore/MK513616 |
| Verona.ITA/19.17/2/D8 | MK513626 | https://www.ncbi.nlm.nih.gov/nuccore/MK513626 |
| Padova.ITA/12.17/1/B3 | MK513599 | https://www.ncbi.nlm.nih.gov/nuccore/MK513599 |
| Padova.ITA/16.17/5/B3 | MK513614 | https://www.ncbi.nlm.nih.gov/nuccore/MK513614 |

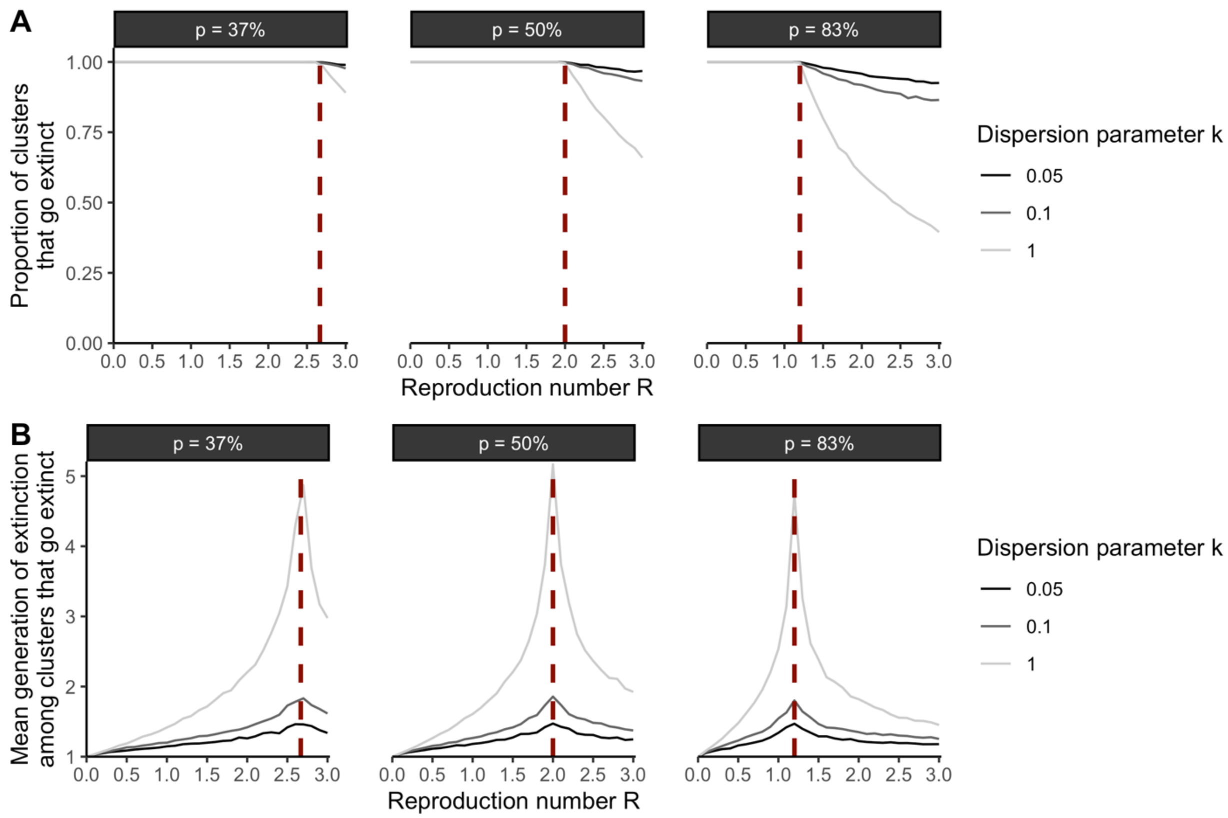


**Figure S1: Dynamics of extinction for clusters of identical pathogen sequences. A.** Proportion of clusters of identical sequences that go extinct as a function of the reproduction number *R* (x-axis) exploring different assumptions regarding the dispersion parameter *k* (colored lines) and the probability p that an infector and an infectee have the same consensus sequence. **B.** Mean number of generations until cluster extinction (among clusters that go extinct) extinct as a function of the reproduction number *R* (x-axis) exploring different assumptions regarding the dispersion parameter *k* (colored lines) and the probability *p* that an infector and an infectee have the same consensus sequence. The vertical red dashed lines correspond to the inverse of the probability *p* that that an infector and an infectee have the same consensus sequence.

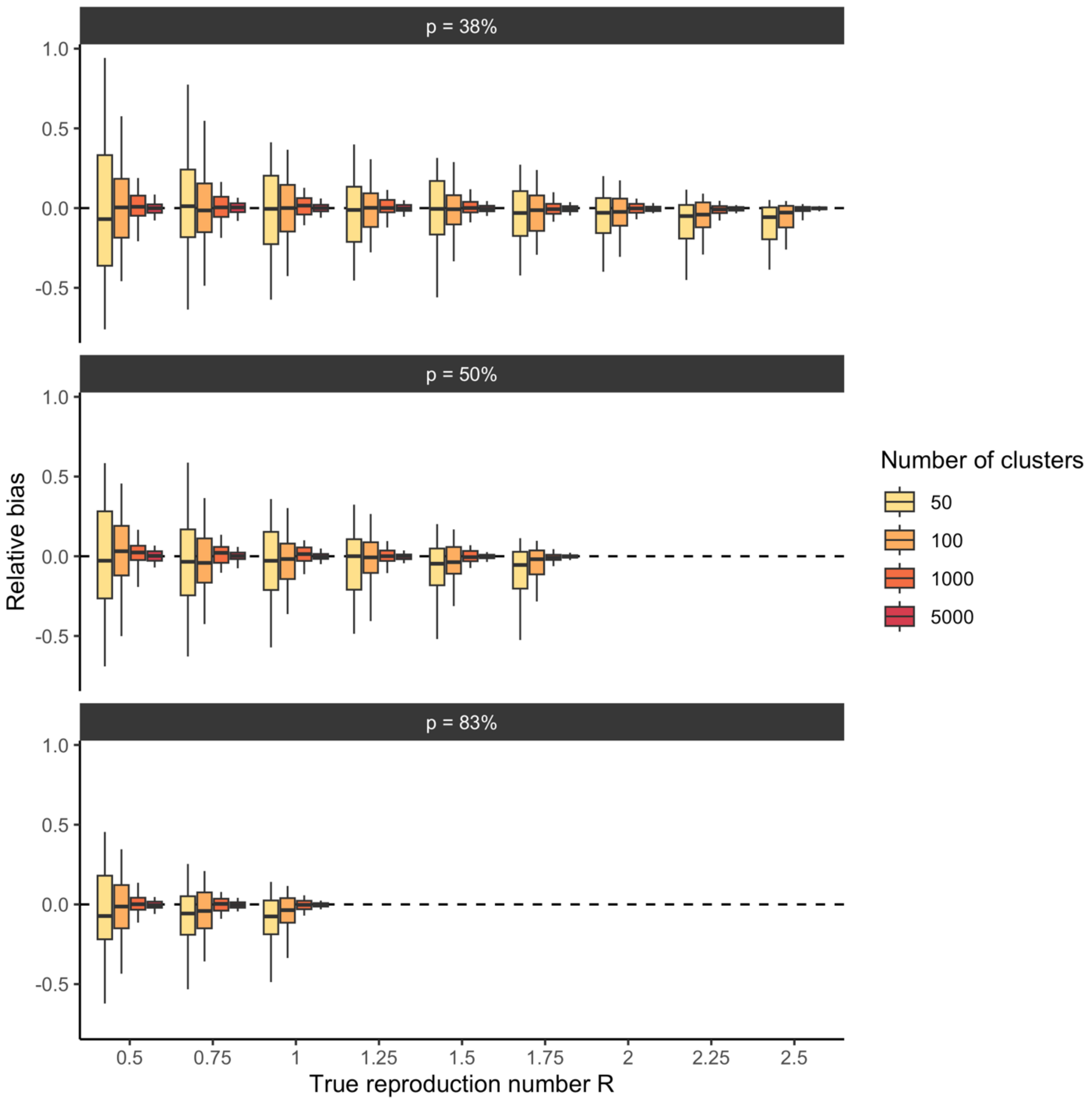


**Figure S2: Relative bias on the reproduction number *R* estimate when the reproduction number lies below the threshold of *1/p*.** For each true value of the reproduction number *R* (x-axis) and value of the probability p that an infector and an infectee have the same consensus sequence, the boxplot depicts the distribution of the relative bias across 100 simulations for different dataset sizes (colours). The relative bias is defined as $(R^{MLE} - R^{true})/R^{true}$ where $R^{true}$ is the true reproduction number used to generate synthetic cluster data and $R^{MLE}$ our maximum likelihood estimates. The simulations were run assuming that 50% of infections were sequenced. The boxplots represent the 2.5%, 25%, 50%, 75% and 97.5% percentiles.

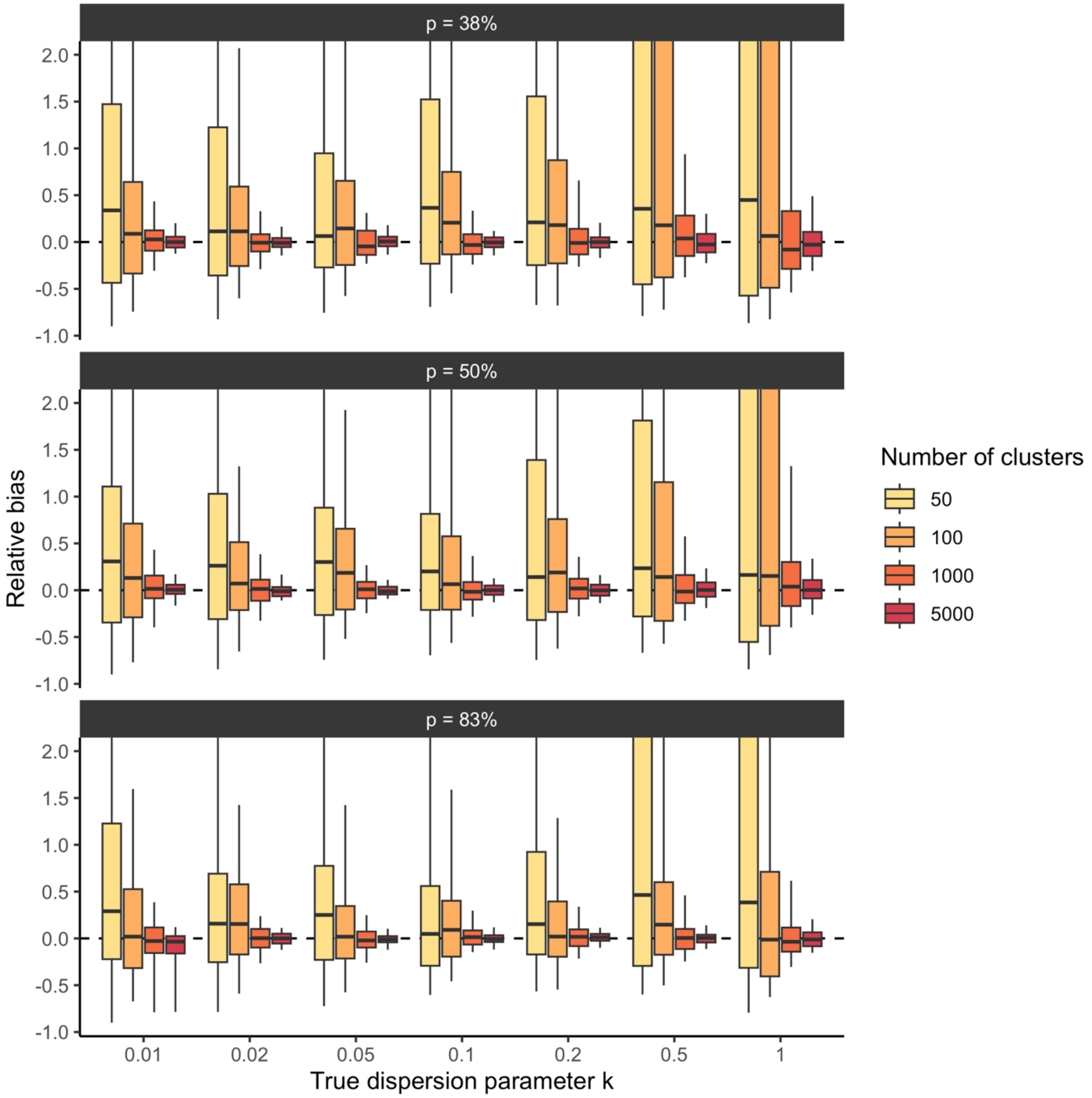


**Figure S3: Relative bias on the dispersion parameter *k* estimate when the reproduction number lies below the threshold of *1/p*.** For each true value of the dispersion parameter *k* (x-axis) and value of the probability *p* that an infector and an infectee have the same consensus sequence, the boxplot depicts the distribution of the relative bias across 100 simulations for different dataset sizes (colours). The relative bias is defined as $(k^{MLE} - k^{true})/k^{true}$ where $k^{true}$ is the true dispersion parameter used to generate synthetic cluster data and $k^{MLE}$ our maximum likelihood estimate. The simulations were run assuming that 50% of infections were sequenced and for a true reproduction number of 1.0. The y-axis was cropped at 2 to increase readability. The boxplots represent the 2.5%, 25%, 50%, 75% and 97.5% percentiles.

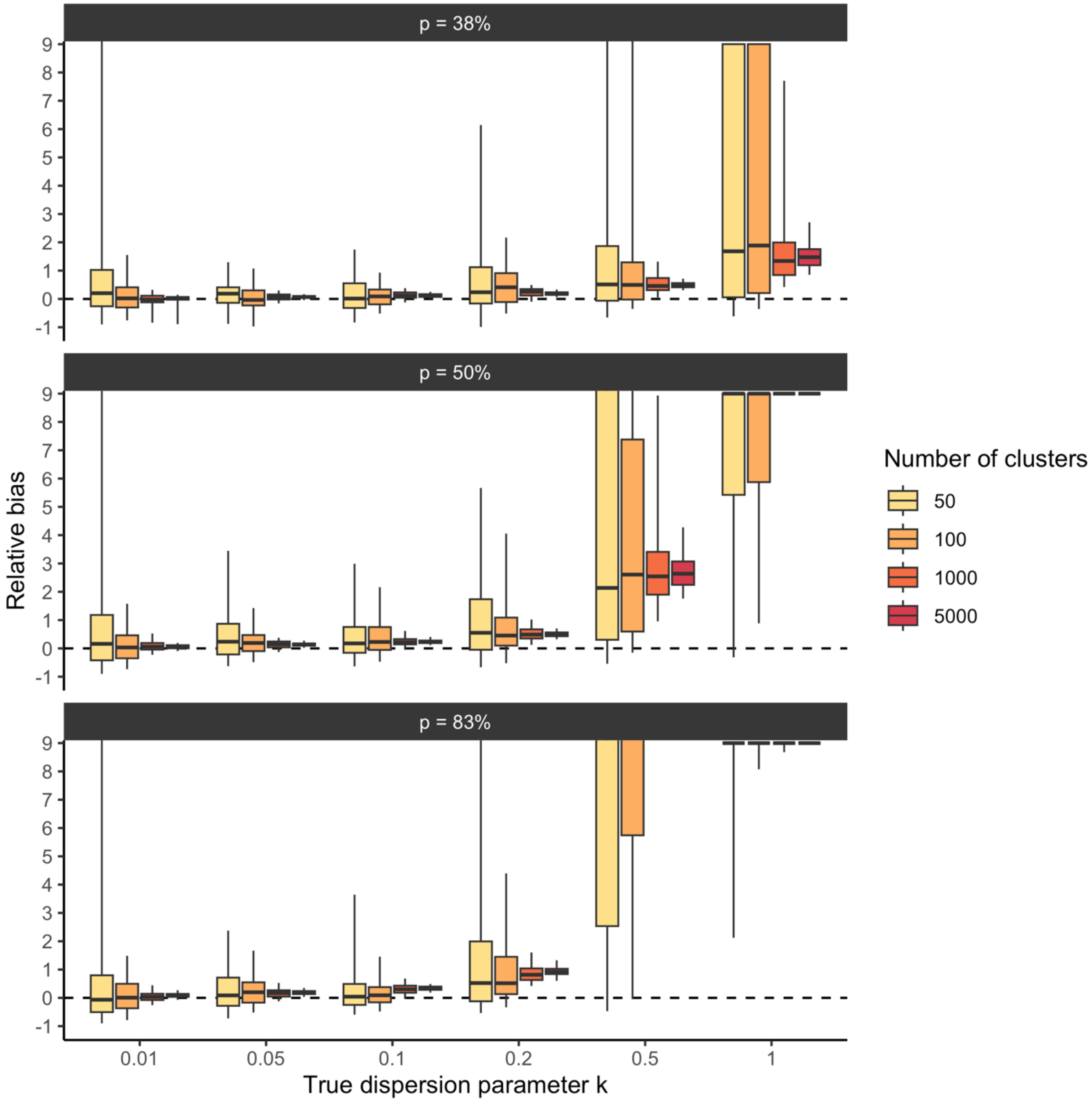


**Figure S4: Relative bias on the dispersion parameter *k* estimate when the reproduction number lies above the threshold of *1/p*.** For each true value of the dispersion parameter *k* (x-axis) and value of the probability *p* that an infector and an infectee have the same consensus sequence, the boxplot depicts the distribution of the relative bias across 100 simulations for different dataset sizes (colours). The relative bias is defined as $(k^{MLE} - k^{true})/k^{true}$ where $k^{true}$ is the true dispersion parameter used to generate synthetic cluster data and $k^{MLE}$ our maximum likelihood estimate. The simulations were run assuming that 50% of infections were sequenced and for a true reproduction number of 3.0. The y-axis was cropped at 9 to increase readability. The boxplots represent the 2.5%, 25%, 50%, 75% and 97.5% percentiles.

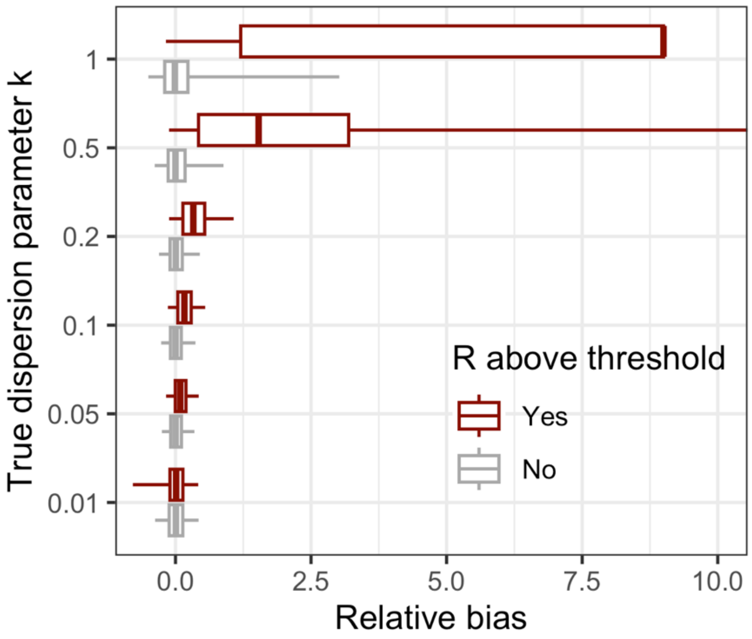


**Figure S5: Impact of reaching the reproduction number threshold on dispersion parameter estimates.** The relative bias is defined as $(k^{MLE} - k^{true})/k^{true}$ where $k^{true}$ is the true dispersion parameter used to generate synthetic cluster data and $k^{MLE}$ our maximum likelihood estimate. The boxplots depict the 2.5%, 25%, 50%, 75% and 97.5% percentiles of relative bias obtained across all the simulations we performed and that are detailed in the methods section.

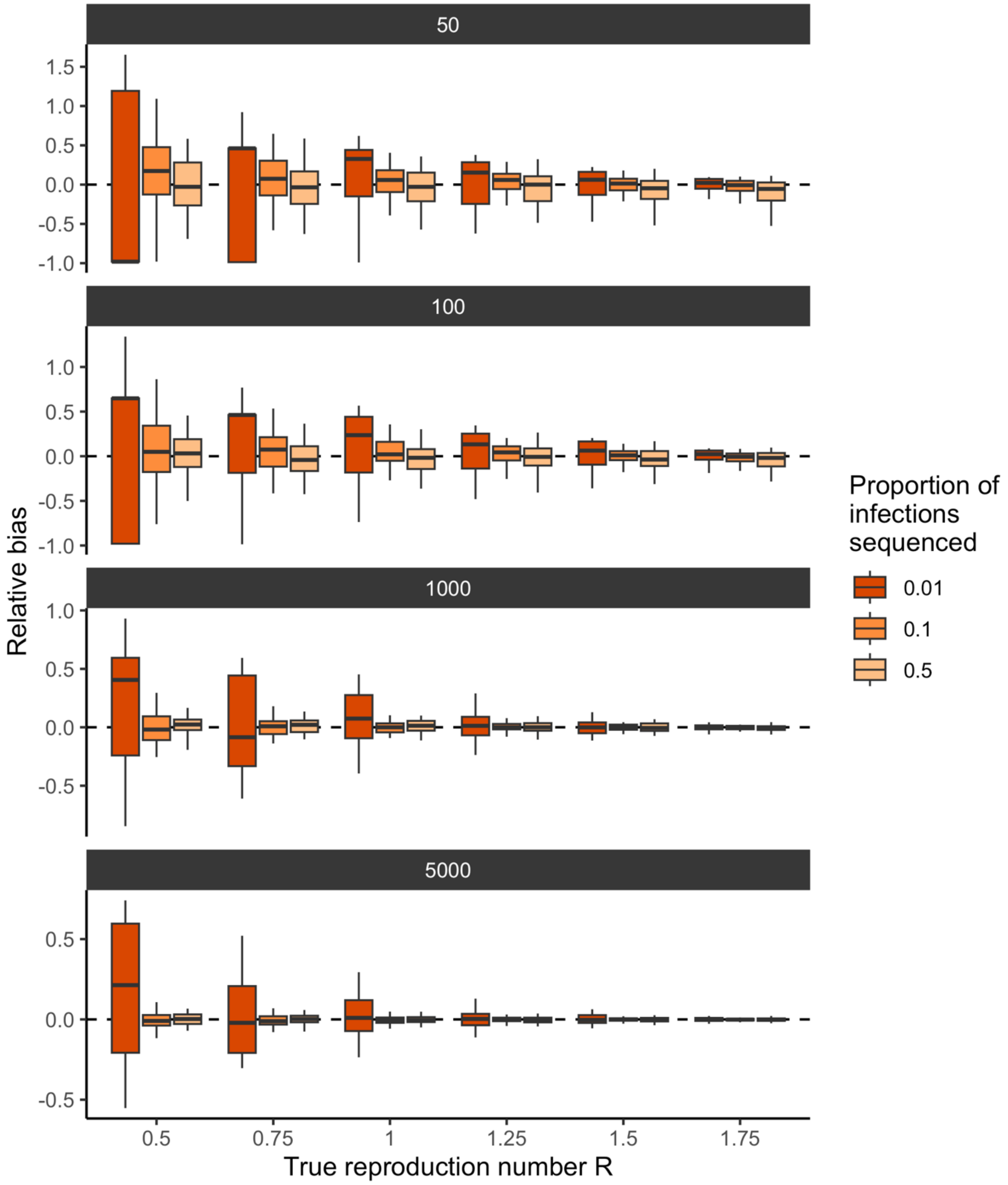


**Figure S6: Impact of the proportion of infections sequenced on the relative bias on the reproduction number *R* estimate.** Results are reported for a probability that an infector and an infectee have the same consensus sequence of 50% and a dispersion parameter value of 0.1. For each true value of the reproduction number *R* (x-axis) and different dataset sizes (different subplots), the boxplots depict the distribution of the relative bias across 100 simulations for different proportion of infections sequenced (colours). The relative bias is defined as $(R^{MLE} - R^{true})/R^{true}$ where $R^{true}$ is the true reproduction number used to generate synthetic cluster data and $R^{MLE}$ our maximum likelihood estimates. The simulations were run assuming that 50% of infections were sequenced. The boxplots represent the 2.5%, 25%, 50%, 75% and 97.5% percentiles.

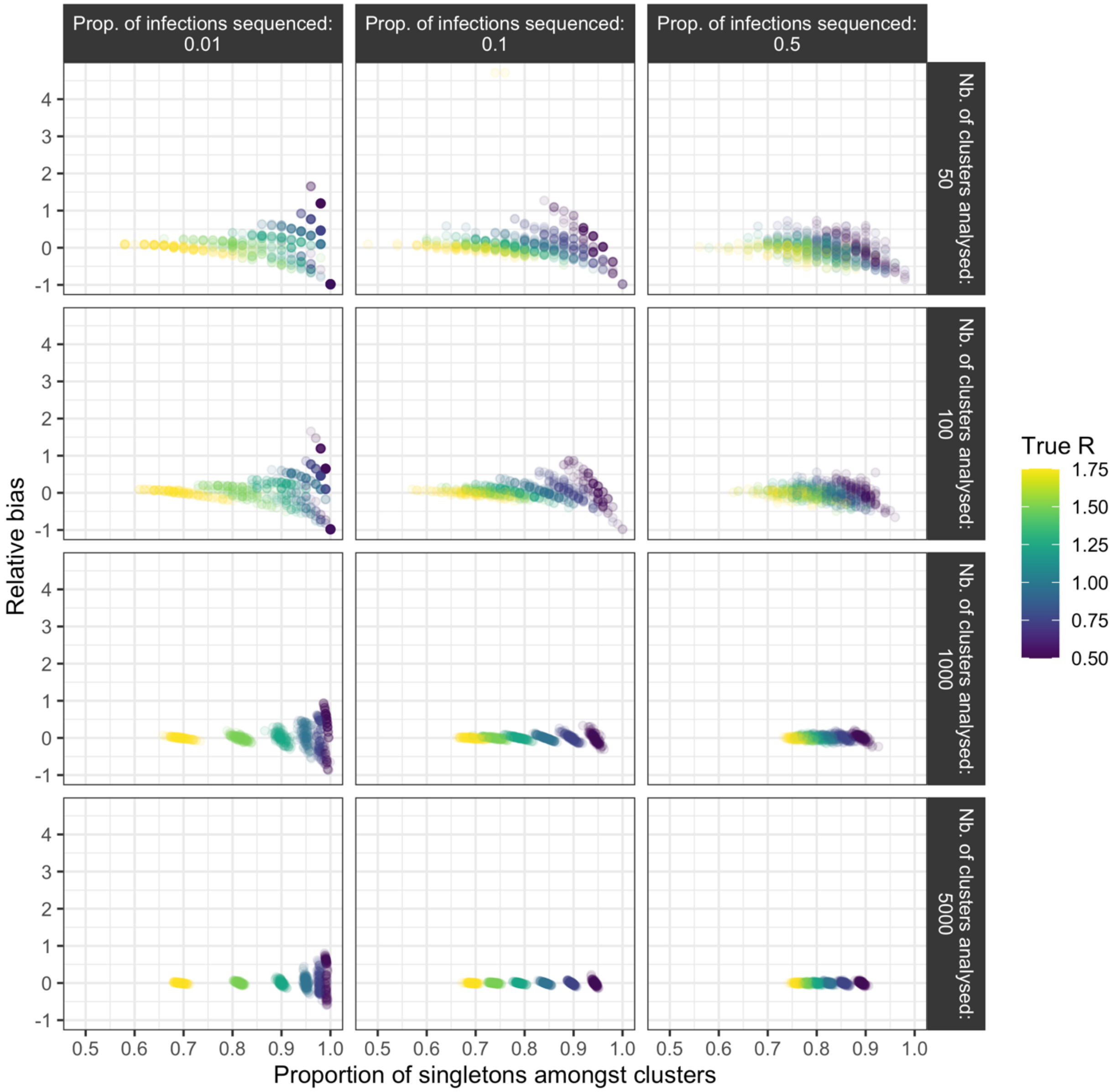


**Figure S7: Relationship between the proportion of singletons among clusters analyzed and the relative bias on the reproduction number *R* estimate.** Different assumptions regarding the proportion of infections sequenced (columns) and the size of the dataset on which the inference was run (rows) are explored. Points are coloured by true reproduction number value. Results are reported for a probability that an infector and an infectee have the same consensus sequence of 50% and a dispersion parameter value of 0.1. The relative bias is defined as $(R^{MLE} - R^{true})/R^{true}$ where $R^{true}$ is the true reproduction number used to generate synthetic cluster data and $R^{MLE}$ our maximum likelihood estimates.

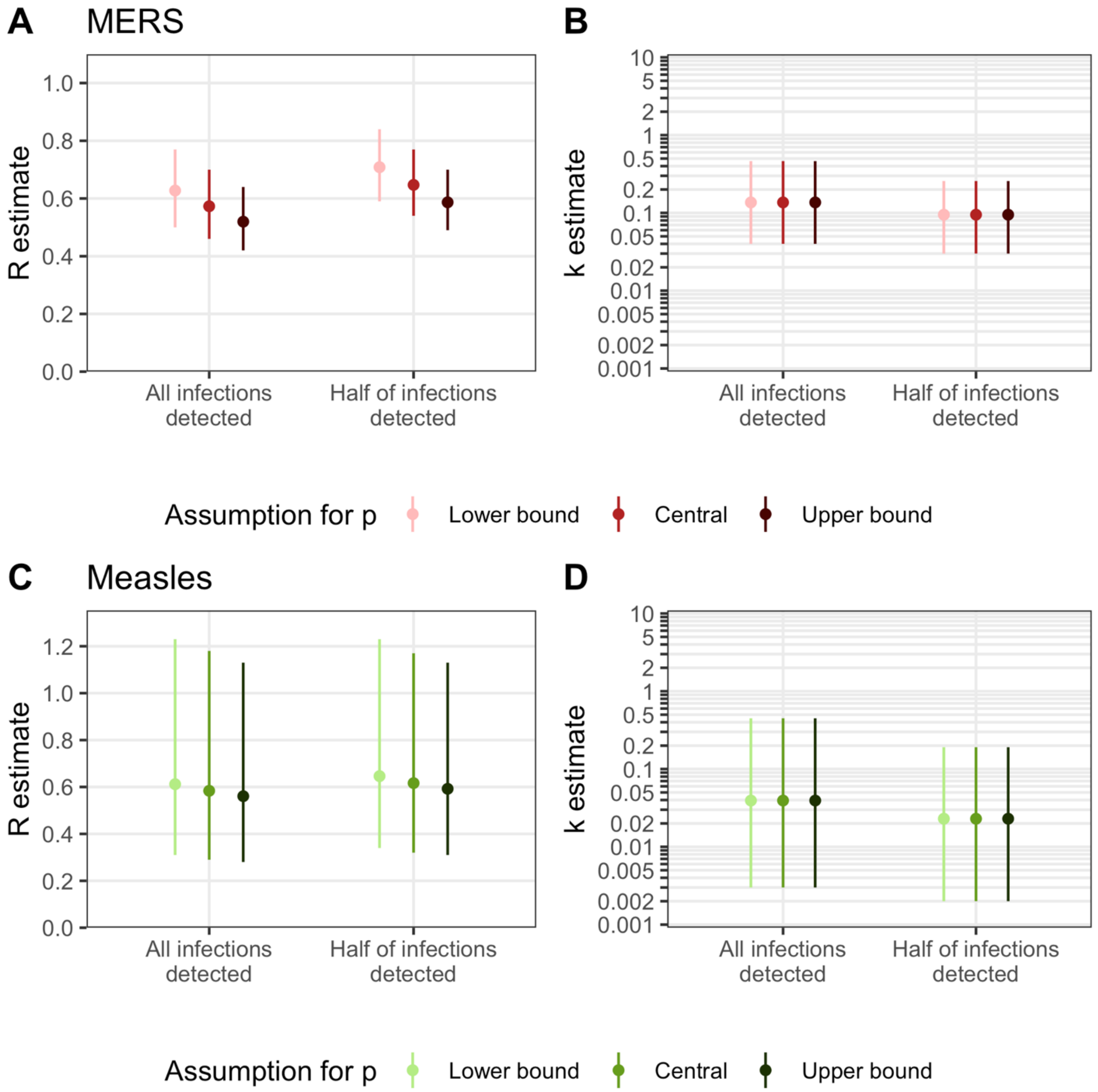


**Figure S8: Sensitivity analysis exploring how *R* and *k* estimates for MERS and measles are impacted by assumptions regarding the probability *p* that an infector and an infectee have the same consensus sequence.** Estimates of **A.** the reproduction number R and **B.** the dispersion parameter *k* for MERS. Estimates of **C.** the reproduction number *R* and **D.** the dispersion parameter *k* for measles during the 2017-2018 Italy outbreak.

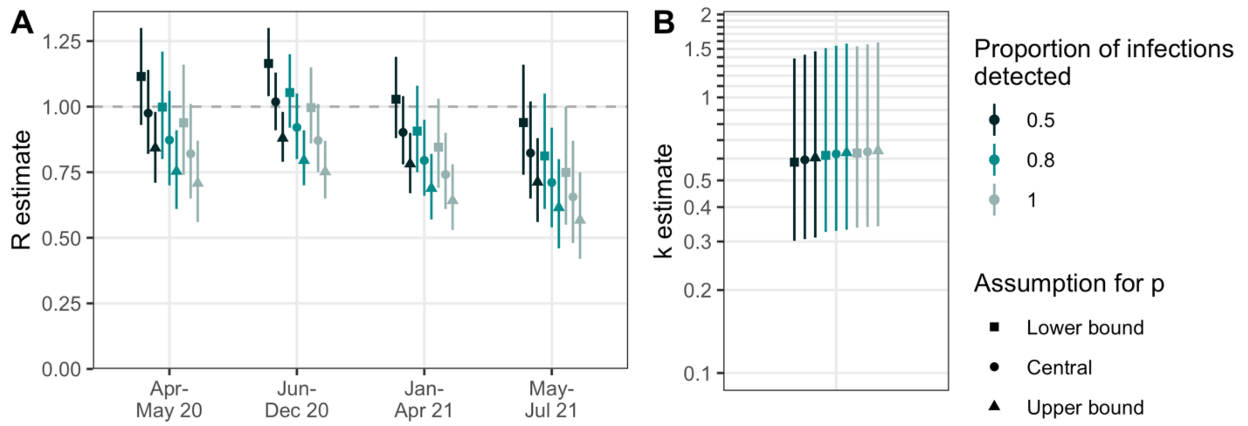


**Figure S9: Sensitivity analysis exploring how *R* and *k* estimates for SARS-CoV-2 in New Zealand are impacted by assumptions regarding the probability p that an infector and an infectee have the same consensus sequence.** Estimates of **A.** the reproduction number *R* and **B.** the dispersion parameter *k* for SARS-CoV-2 in New Zealand.

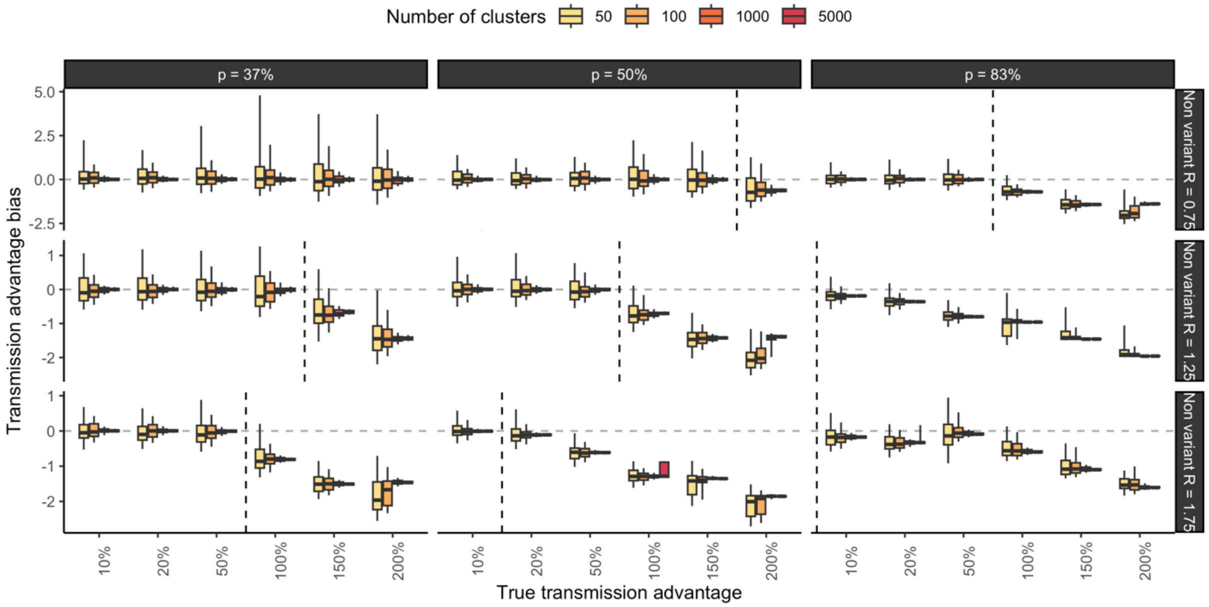


**Figure S10: Transmission advantage bias as a function of the true transmission advantage and varying the probability that an infector and an infectee have the same consensus sequence (rows) and the reproduction number of the non-variant $R_{NV}$ (columns).** Each subplot corresponds to a given assumption regarding the probability that an infector and an infectee have the same consensus sequence and the reproduction number of the non-variant. In each subplot, the vertical dashed line corresponds to the limit from which the reproduction number of the variant $R_V$ reaches the threshold of *1/p*. Vertical dashed lines before the 10% x-axis tick correspond to situations where the reproduction number of the non-variant $R_{NV}$ is also above the threshold of *1/p*.

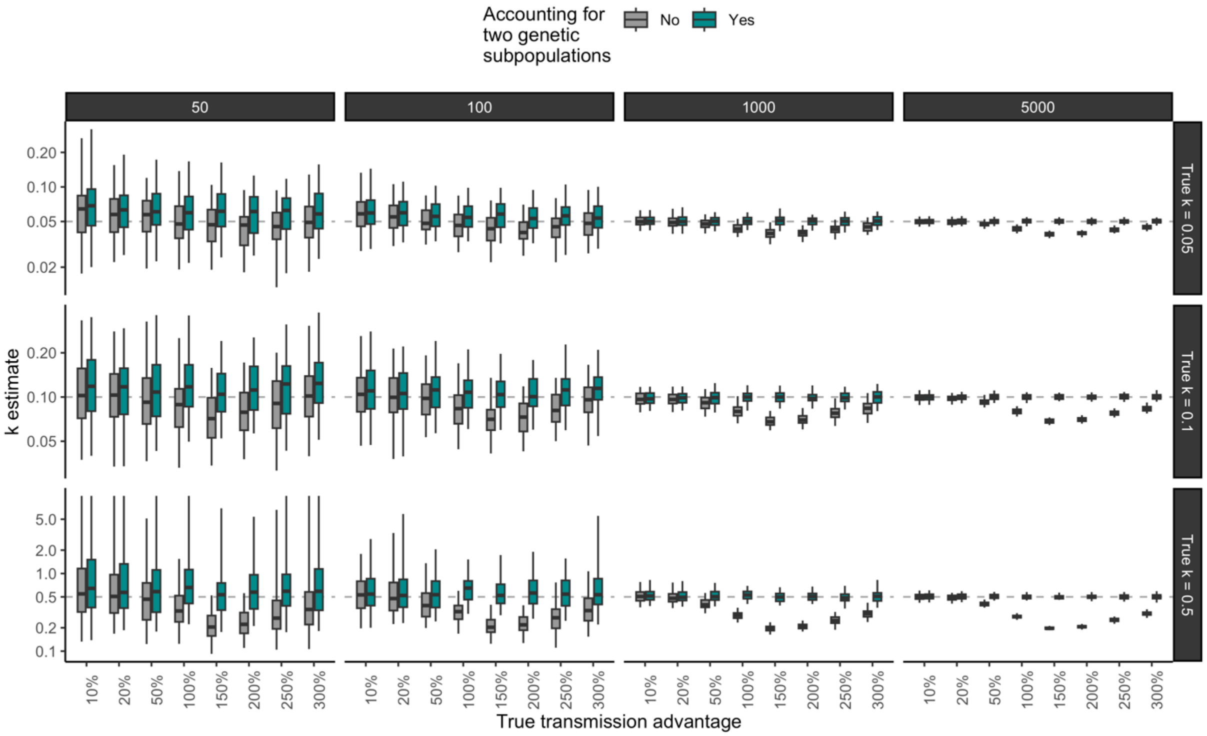


**Figure S11: Impact of accounting for different genetic subpopulations on estimates of the dispersion parameter for different assumptions regarding the true dispersion parameter (rows) and different dataset sizes (columns)** In each subplot, the horizontal dashed grey line corresponds to the true dispersion parameter value used to generate synthetic clusters of identical sequences. The boxplots summarize the 2.5%, 25%, 50%, 75% and 97.5% percentile of maximum-likelihood estimates obtained across 100 simulated datasets.

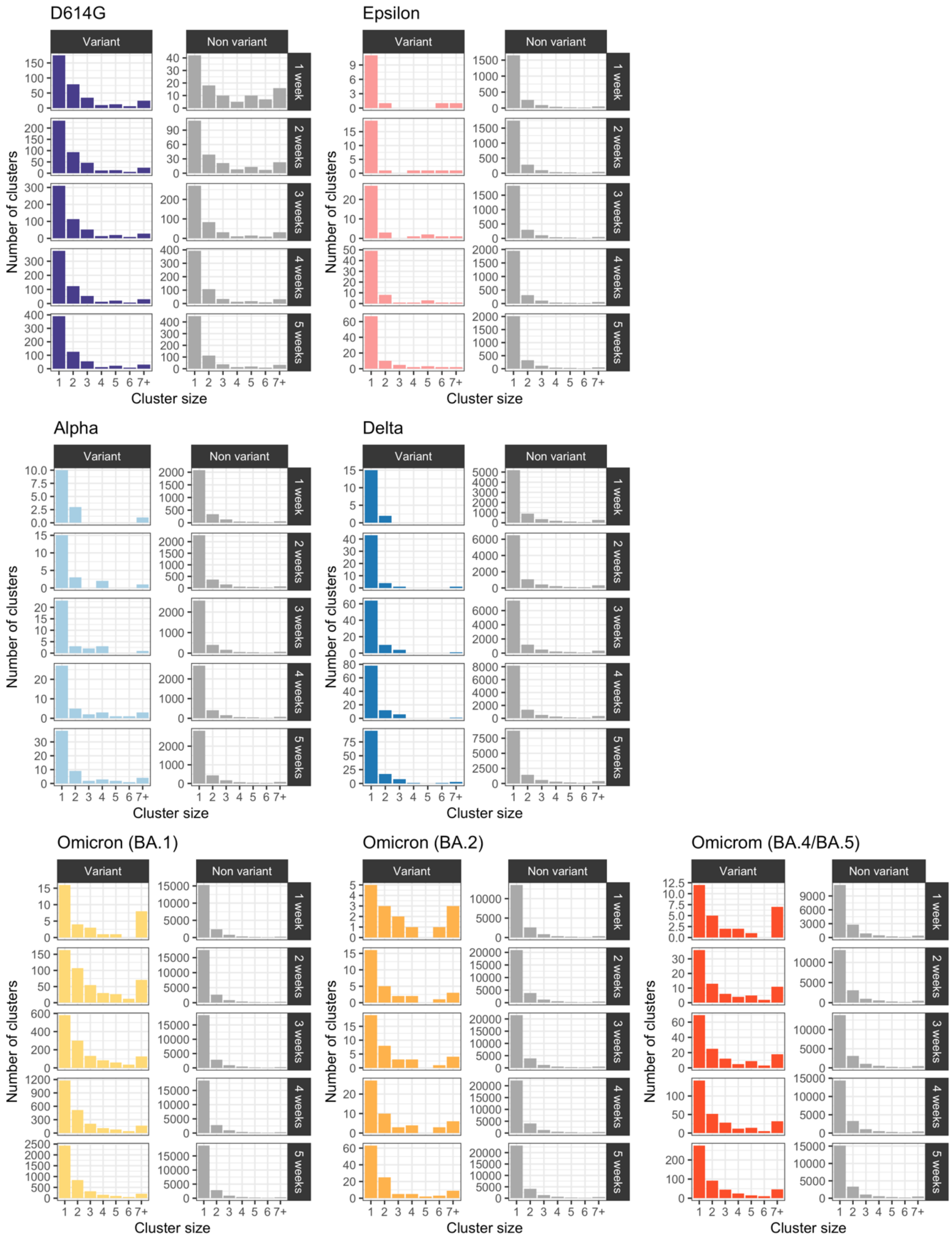


**Figure S12: Size distribution of clusters of identical SARS-CoV-2 sequences in Washington state split by variant of interest.** For each variant that we studied, we displayed the distribution of cluster sizes for the variant and the non-variant considering different time window length (1 to 5 weeks). The time windows begin when the cumulative number of collected in Washington state variant sequences reached 10 (See Table S8). We considered that clusters of identical sequences fell into the time-window if they were first detected during that time window.

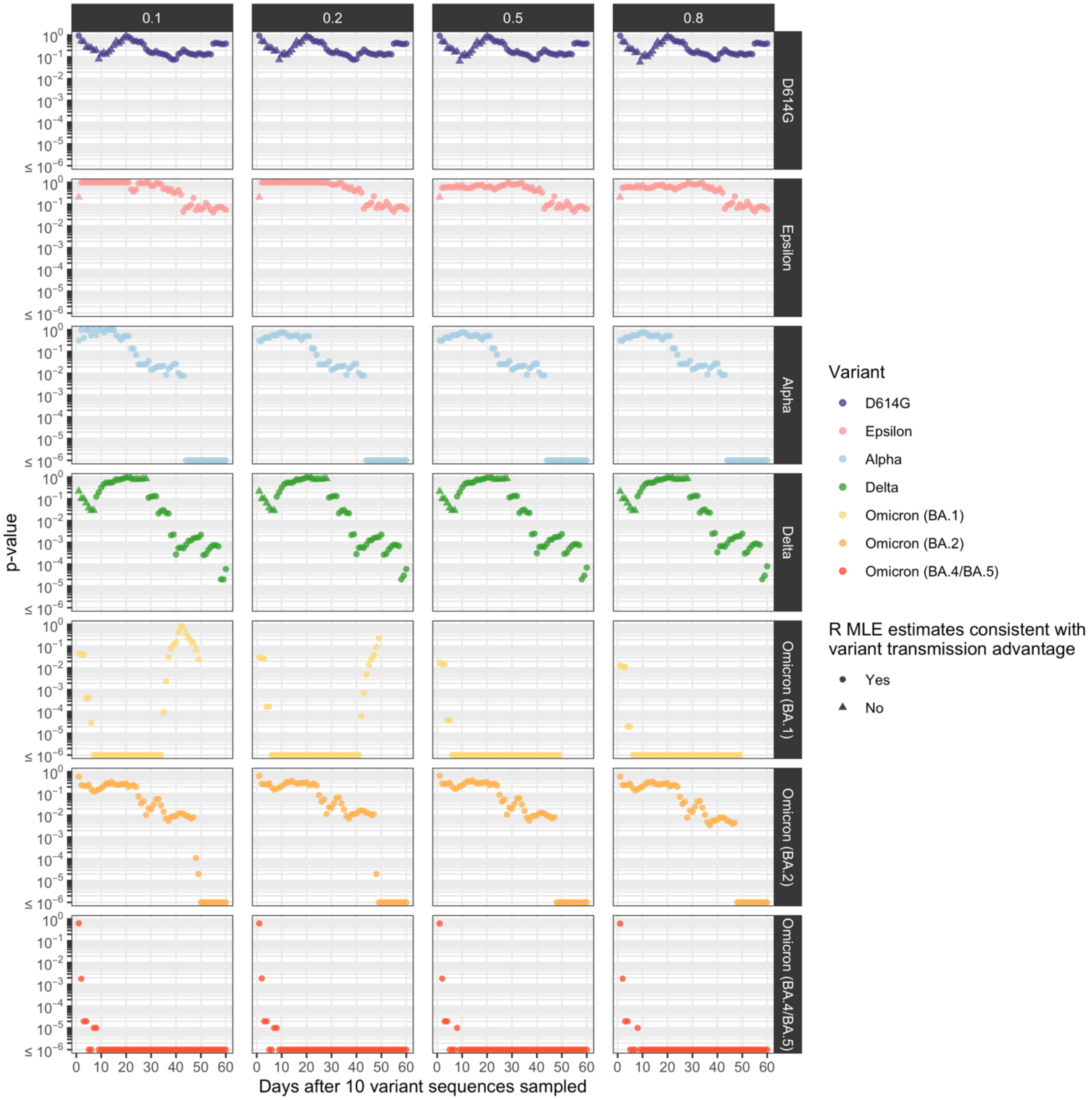


**Figure S13: Sensitivity analysis varying our assumption regarding the fraction of infection detected as cases (different panels) on the p-values for variant transmission advantage in WA state.** P-values over time since collection of 10 variant sequences for different SARS-CoV-2 variants during the COVID-19 pandemic in Washington state exploring different assumptions regarding the fraction of infections detected (columns). We considered maximum likelihood estimated (MLE) to be consistent with a variant transmission advantage if the estimated reproduction number of the variant was higher than that of the non-variant.

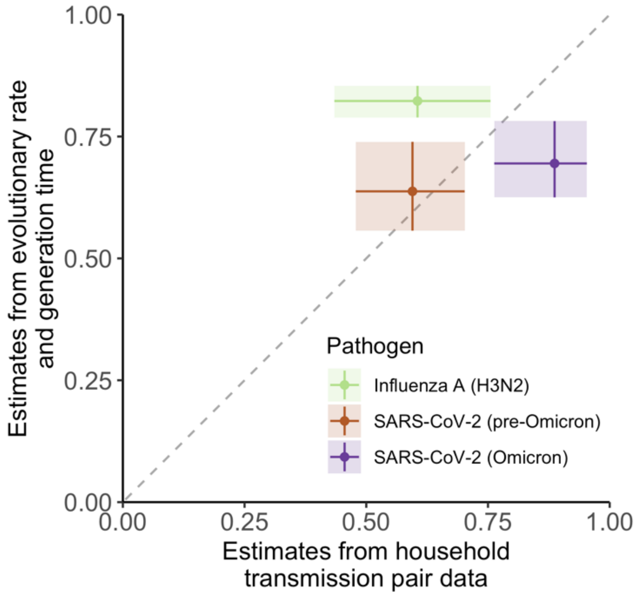


**Figure S14: Comparison between estimates of *p* obtained from household transmission pair data and from assumptions regarding the evolutionary rate and the generation time.** Vertical segments correspond to our uncertainty ranges around *p* estimates. Horizontal segments correspond to 95% binomial confidence intervals around the proportions obtained from transmission pair data.

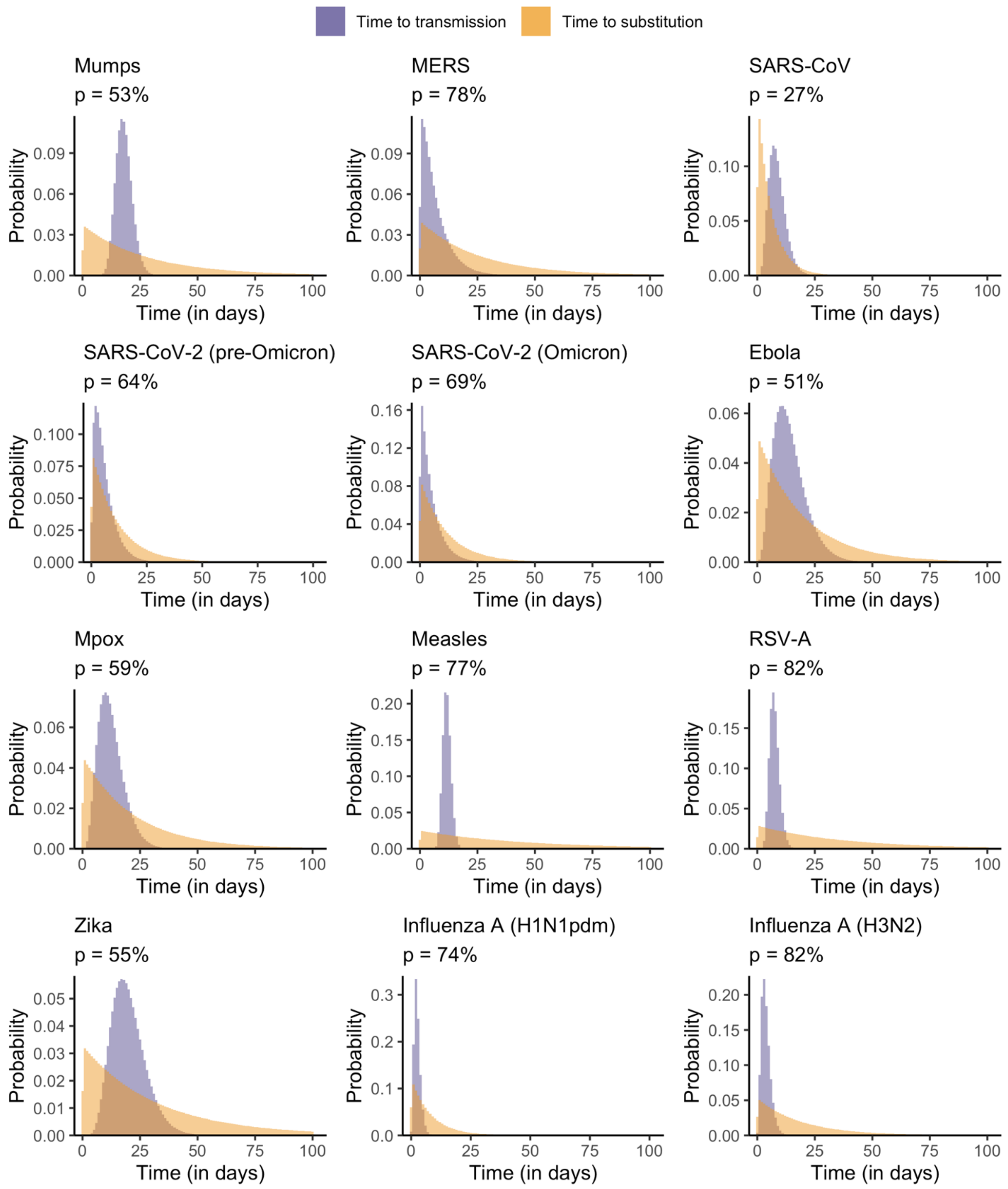


**Figure S15: Comparison of the distribution of the time to occurrence of a first substitution and the time to transmission for different pathogens**. For each pathogen, we additionally report the estimated probability *p* that transmission occurs before substitution. Here, we depict the simulations corresponding to the central estimate for the substitution rate and the generation time.

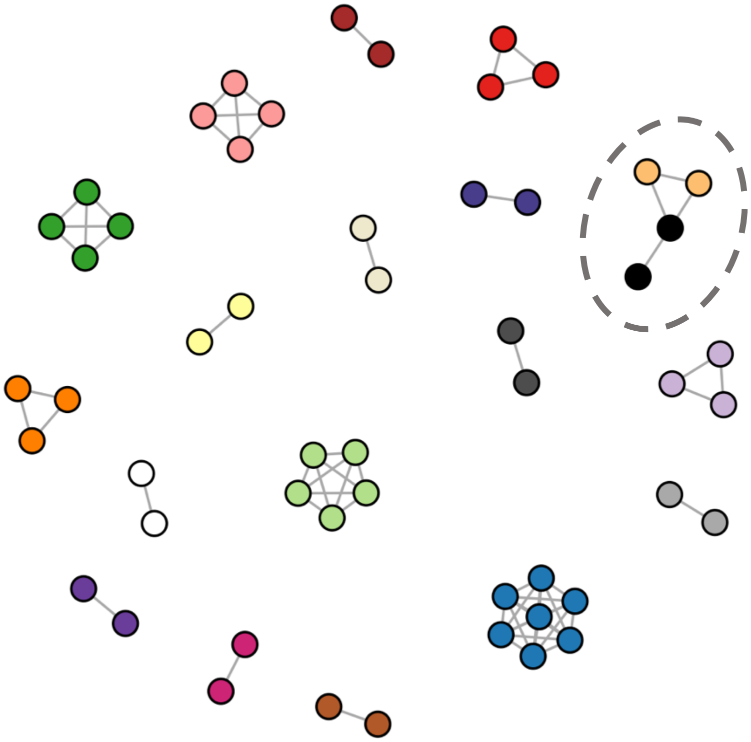

**Figure S16: Difference between identical sequences obtained from the distance matrix and the reconstructed clusters of identical sequences for MERS-CoV sequences.** Each vertex corresponds to a MERS-CoV sequence. Vertices are connected if their pairwise distance is equal to 0. Vertices have the same colour if they were allocated to the same cluster of identical sequences. The clusters for which there is a disagreement between the distance matrix and the cluster allocation (i.e. when some identical sequences are not in the same cluster) are circled. For clarity, we only displayed sequences with at least one other identical sequence in the pairwise distance matrix.

**Supplementary text A – Impact of infectious duration and transmission bottleneck size on the proportion of transmission pairs with identical consensus sequences**

In this manuscript, we assumed that the proportion of infectees that have the same consensus sequence as their infector could be approximated by the probability that a transmission event occurs before a substitution event. In this section, we report a simulation exercise that was conducted to evaluate the relevance of this assumption. In our simulations, we explore different assumptions regarding the transmission bottleneck size and the duration of infectiousness. We do not account for within-host selection or selection at transmission. This assumption lies in line with studies showing that the within-host diversity of several RNA viruses causing acute infections (the class of pathogen for which we developed our novel methods) is mainly shaped by purifying selection and neutral evolution (32, 33). The simulations account for changes in population size over time by accounting for births and deaths and assuming that the pathogen population grows to an equilibrium within-host population size. However, we do not account for changes in population size that might arise from within-host selective processes.

*Description of the simulations*

We used the SEEDY R package developed by Worby and Read (34) to simulate deep-sequencing in transmission pairs under different assumptions regarding the transmission bottleneck size (1, 2, and 10) and the generation time (6, 12, 100 or 200 days). We used these transmission pairs to compute the fraction of pairs that had the same consensus genome and compared this with the probability that a transmission event occurs before a substitution one.

We considered the spread of a pathogen with a genome of length 10,000 kb. We assumed a pathogen equilibrium within-host population size of 1000 and that the pathogen would undergo 3 generations per day. We assumed that substitutions occur on average every 12 days. We assumed that individuals were sequenced at a random time between when they were infected and when they infect the recipient. Recipients are sampled at random between when they are infected and the end of their infectious period. For each scenario, we generated 1200 transmission pairs.

*Within-host population dynamics*

We used the simulation framework implemented in the SEEDY R package (34) to incorporate pathogen within-host population dynamics. Let $N_{eq}$ denote the within-host equilibrium population size and $N_t$ denote the pathogen within-host population size at time $t$. The within-host population is seeded at $N_B$ (transmission bottleneck population size) and grows to a size $N_{eq}$. Per-generation deaths occur with a probability of $N_t / (2N_{eq})$. Let $N_t^g$ denote the within-host population size at time $t$ of a genotype $g$. $N_{t+1}^g$ is then drawn from a binomial distribution as follows:

$$N_{t+1}^{g} \sim B(N_t^g, N_{t-1}/N_{eq})$$

*Relationship between frequency of a mutant in the donor (infector) and the recipient (infectee)*

Figure S17 depicts the relationship between the frequency of a mutant allele in the donor of a transmission pair and the recipient. As transmission bottleneck size increases, we observe more points inside the unity square (square between (0,0) (0,1) (1,1) and (1,0)). This is consistent with more intra-host variant being passed from the donor to the recipient.

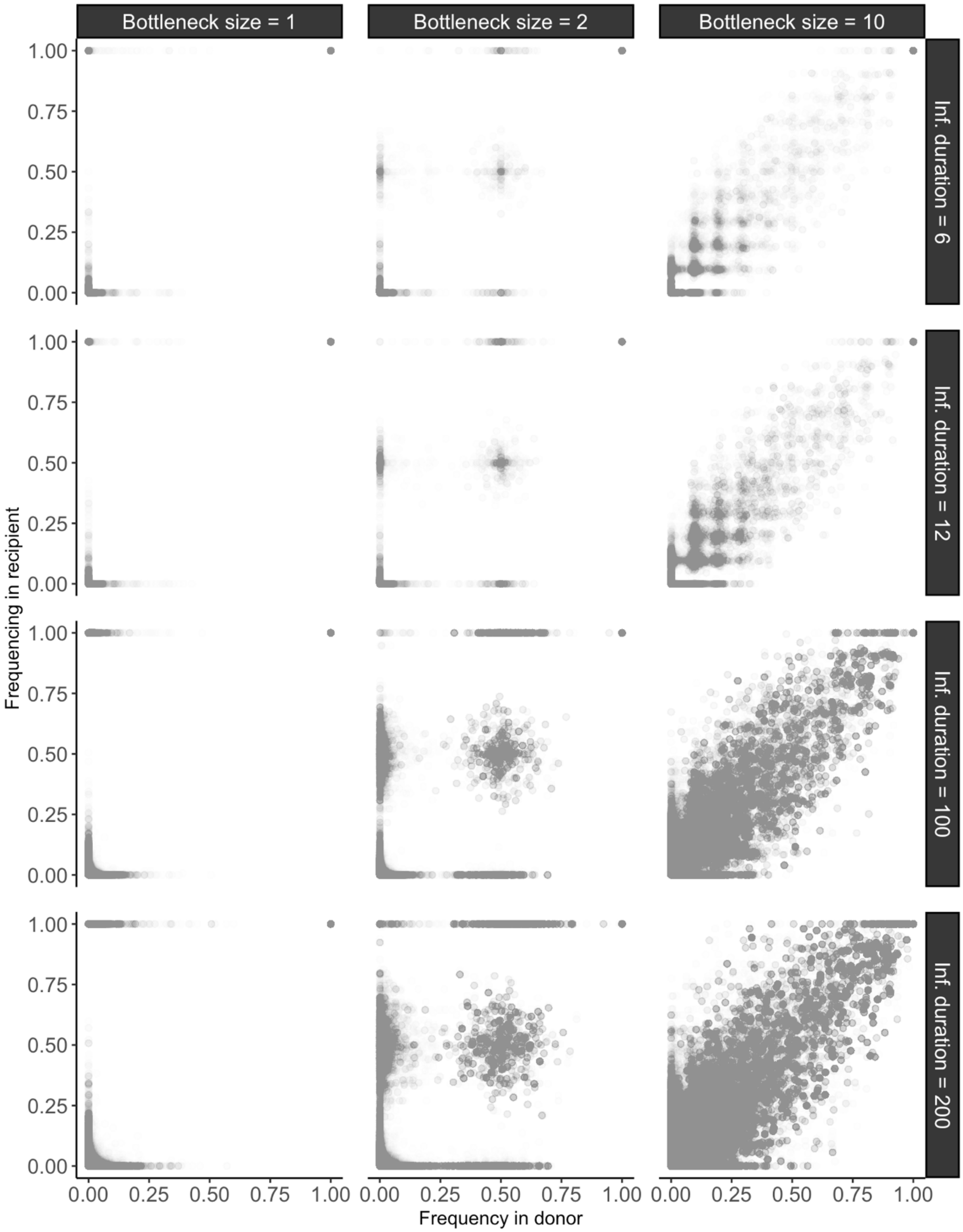


**Figure S17: Relationship between the frequency of a mutant in the donor (infector) and the recipient (infectee) of a transmission pair.** Different assumptions regarding the transmission bottleneck size (different columns) and the generation time (in days – different rows) are explored.

*Proportion of transmission pairs with identical consensus genomes from the simulations*

For each of these scenarios, we computed the fraction of transmission pairs with identical consensus sequences and compared this to the theoretical probability that a transmission event occurs before a substitution one (dashed horizontal lines in Figure S18). For narrow transmission bottlenecks of size 1, the theoretical probability value approximates well the proportion of pairs with identical consensus sequences. As transmission bottleneck size widens, this theoretical probability no longer approximates well the proportion of pairs with identical consensus sequences. When simulating the spread and evolution of a pathogen characterized by a longer generation time (200 days), we found that the proportion of transmission pairs with identical consensus genomes differed slightly from that expected from the theoretical probability that a transmission event occurs before a substitution event.

To conclude, this simulation study shows that the probability that transmission occurs before substitution is a good proxy for the proportion of pairs with identical consensus genomes for pathogens characterized by narrow transmission bottlenecks, relatively short infectious durations and limited within host-diversity.

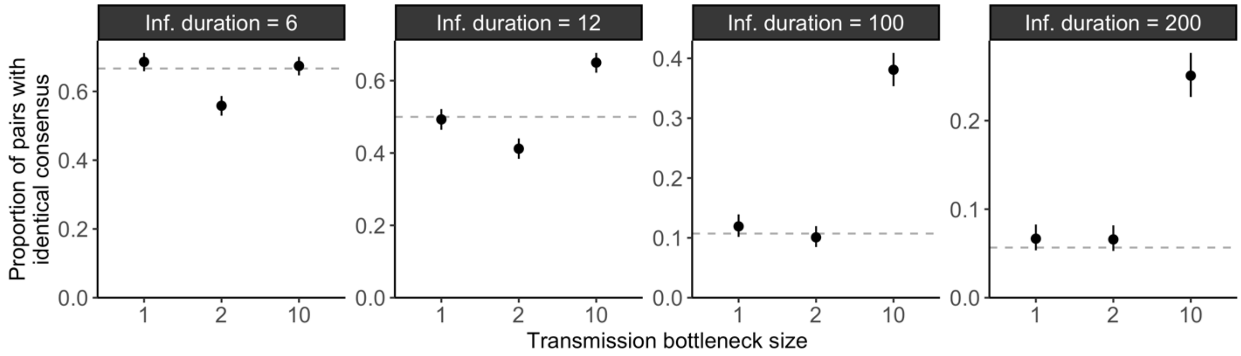


**Figure S18: Proportion of transmission pairs with identical consensus sequences** exploring different assumptions regarding the transmission bottleneck size and the disease generation time ("inf. duration" in days). Vertical segments correspond to 95% confidence intervals. Each point was obtained by generating 1,200 transmission pairs. The horizontal dotted lines correspond to the probability that a transmission event occurs before a substituion event.

*Within-host diversity*

Our simulation framework makes a number of simplifying assumptions regarding the within-host mutation process and the pathogen's population dynamics. We checked whether our simulations were associated with reasonable outputs in terms of within-host diversity. We computed the nucleotide diversity as the mean number of single nucleotide polymorphism (SNP) differences per site across scenarios (Figure S19).

As expected, we found that within-host pairwise nucleotide diversity increases with both the duration of infectiousness and the size of the transmission bottleneck. For pathogens characterized by narrow transmission bottleneck (1 to 2) and short generation times, we obtained nucleotide diversity at the genome level of the order of $10^{-5}$ to $10^{-4}$ SNPs per site. This fits within the range of estimates obtained across RNA viruses causing acute infections ($10^{-6}$ to $10^{-4}$ for SARS-CoV-2 (33, 35), $10^{-5}$ for RSV (36, 37), $10^{-4}$ for DENV-1 (38), $10^{-5}$ for influenza A viruses (39)).

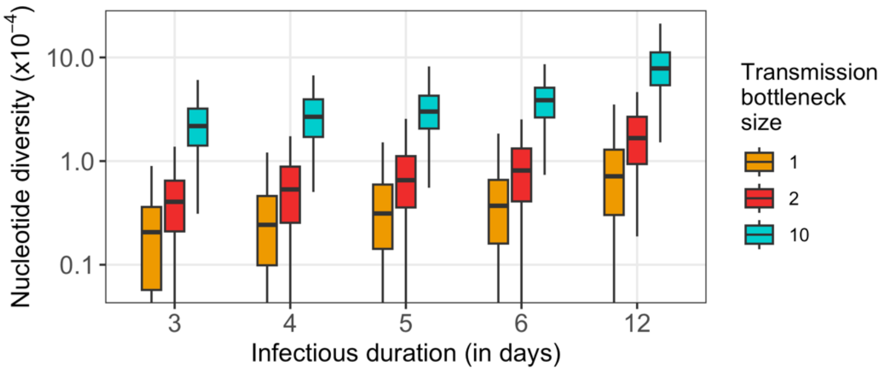


**Figure S19: Nucleotide diversity for different infectious durations and transmission bottleneck size.** Nucleotide diversity was computed as the mean within-host SNP difference per site. Boxplots indicate the 2.5%, 25%, 50%, 75% and 97.5% quantiles. Vertical whiskers going to the bottom of the plot correspond to 0 values.

**Supplementary text B - Inference of transmission parameters conditional on cluster extinction**

In the main text, we showed that our inference framework provides unbiased estimates of both R and k when the mean number of offspring with identical sequences is lower than 1 ($R \cdot p < 1$), corresponding to situations where cluster extinction is almost certain. Our method however becomes unreliable when the probability of cluster extinction is strictly lower than 1. This was done relying on the full distribution of clusters of identical sequences, including those that did not go extinct (which were then set to an arbitrary high threshold value). An alternative approach would consist in looking at cluster sizes conditional on extinction (40, 41). Previous theoretical work has indeed shown that a supercritical epidemic process where extinction is uncertain (characterized by *R > 1*) can be mapped to a subcritical counterpart characterized by a mean number of offspring lower than 1 (*R < 1*) and the same dispersion parameter (40).

In the following paragraphs, we show how our inference framework could be adapted to look at the size of clusters of identical sequences conditional on them having gone extinct. We then evaluate the performance of this adapted statistical framework and highlight some remaining challenges for real-world applications.

*Distribution of the size of clusters of identical sequences conditional on extinction*

We used the formalism introduced by Waxman and Nouvellet (40) to describe the size distribution of clusters of identical pathogen sequences conditional on extinction. They showed that, conditional on extinction, supercritical and subcritical dynamics (respectively characterized by a reproduction number below and above 1) cannot be distinguished. Waxman and Nouvellet had characterized the size of finite disease outbreaks. Here, we instead consider finite *mutation-less* outbreaks, *i.e.* clusters of infected individuals characterized by the same pathogen sequence.

Let $\epsilon$ denote the probability of extinction for a cluster of identical pathogen sequences. If the mean number of offspring with identical sequences is lower than 1, $\epsilon$ is equal to 1. Otherwise, $\epsilon$ is lower than 1. Following Waxman and Nouvellet, we introduce $R_s$, as the reproduction number associated with clusters of identical sequences that got extinct. In the following, we refer to $R_s$ as the subcritical reproduction number. We have the following relationship between the reproduction number $R$ and $R_s$:

$$R_s = R \cdot \epsilon^{1+\frac{1}{k}} \qquad (*)$$

where $k$ is the dispersion parameter of the offspring distribution. Figure S20 depicts how the subcritical reproduction number $R_s$ is impacted by the reproduction number $R$, the dispersion parameter $k$ and the probability that an infector and an infectee have the same consensus sequence. As Waxman and Nouvellet note, the subcritical reproduction number $R_s$ mirrors the supercritical one $R$. This means that inferring the subcritical reproduction number $R_s$ enables to infer the reproduction number $R$ as there is a direct relationship between the two (Figure S20).

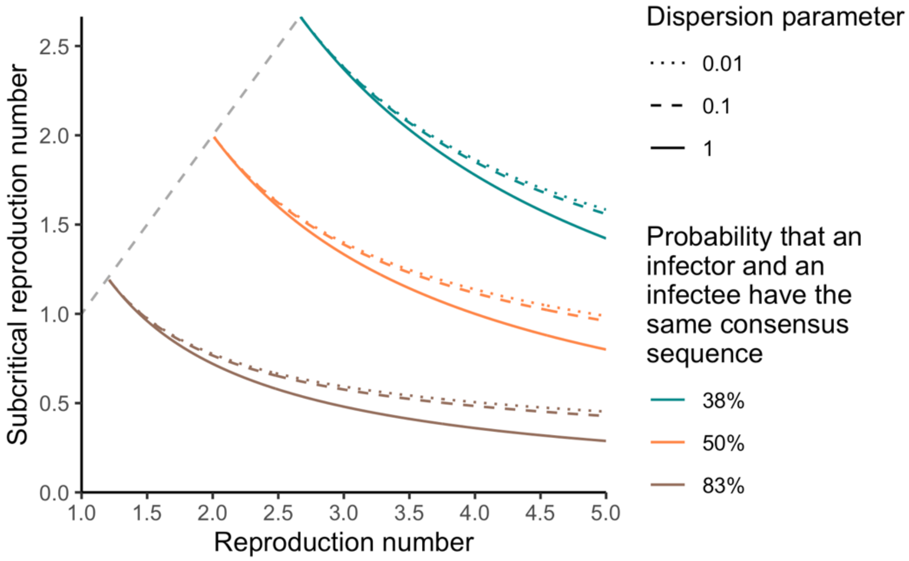


**Figure S20: Relationship between the reproduction number *R* and the subcritical reproduction number *R<sub>s</sub>*** for different probabilities *p* that an infector and an infectee have the same consensus sequence and different values of the dispersion parameter *k*. Colored lines correspond to reproduction numbers lying above the threshold of *1/p*. The dashed grey lines correspond to reproduction numbers lying below the reproduction number threshold (for which the reproduction number is equal to the subcritical reproduction number).

The probability $r_j^{extinct}$ for a cluster of identical sequences to be of size $j$ conditional on extinction is equal to (40, 41):

$$r_j^{extinct} = \frac{\Gamma(kj + j - 1)}{\Gamma(kj) \cdot \Gamma(j + 1)} \cdot \frac{\left(\frac{pR_s}{k}\right)^{j-1}}{\left(1 + \frac{pR_s}{k}\right)^{kj+j-1}}$$

and the probability $r_j$ for a cluster of identical sequences of being of size $j$ is thus equal to:

$$r_j = r_j^{extinct} \cdot \epsilon \qquad (**)$$

More specifically, we note that $R_s < 1/p$. In the specific situation where $R < 1/p$, we have $R = R_s$ and $r_j^{extinct} = r_j$.

*Inference from the size of clusters of identical sequences conditional on extinction*

Assuming we have a dataset comprised of the size of clusters of identical sequences that got extinct, we can hence infer the value of the subcritical reproduction number $R_s$ and the dispersion parameter by using the updated formula $(**)$ for the probability of cluster of identical sequences of being of size $j$ in the derivation of the likelihood. Implementing this updated framework, we then obtained maximum likelihood estimates of $R_s$ and $k$ by imposing values of the reproduction number ranging between 0.01 and $1/p$ and values of the dispersion parameter ranging between 0.001 and 10.0.

We evaluated our inference framework on synthetic cluster data generated using a branching process with substitution (see main text). Clusters were simulated until reaching a maximum size of 10,000. We then considered that clusters who reached 10,000 had not gone extinct and applied our inference framework to the subset that got extinct (size < 10,000). Figure S21-S23 shows that we were able to recover the expected value of the dispersion parameter and the subcritical reproduction number $R_s$.

Assuming prior knowledge on whether the reproduction number lies above or below the threshold of $1/p$, the estimated subcritical reproduction number can either directly be interpreted as the reproduction number of the outbreak (below the threshold) or mapped to a corresponding reproduction number higher than $1/p$ (using equation $(*)$, Figure S20)

*Challenges for the application to real-world data*

In the previous paragraphs, we introduced an alternative approach to characterize the disease offspring distribution when the mean number of offspring with identical genomes is higher than 1. By restricting the analysis to clusters of identical sequences that got extinct, we showed that we could accurately infer the reproduction number and the dispersion parameter.

However, we acknowledge that determining in practice whether a cluster of identical sequences has become extinct may be challenging. Furthermore, we assumed here that the epidemiological process under study was stationary (*i.e.* that the reproduction number and the dispersion parameter are constant throughout the study period). In practice, behaviour changes, the implementation of interventions or the depletion of the susceptible population as the epidemic progresses can modify the effective reproduction number. This is likely especially problematic for reproduction numbers greater than 1 (and by extension above the threshold of $1/p$). Overall, further work is warranted to estimate the offspring distribution's parameters above the threshold of $1/p$ from real-world data describing the size of clusters of identical sequences.

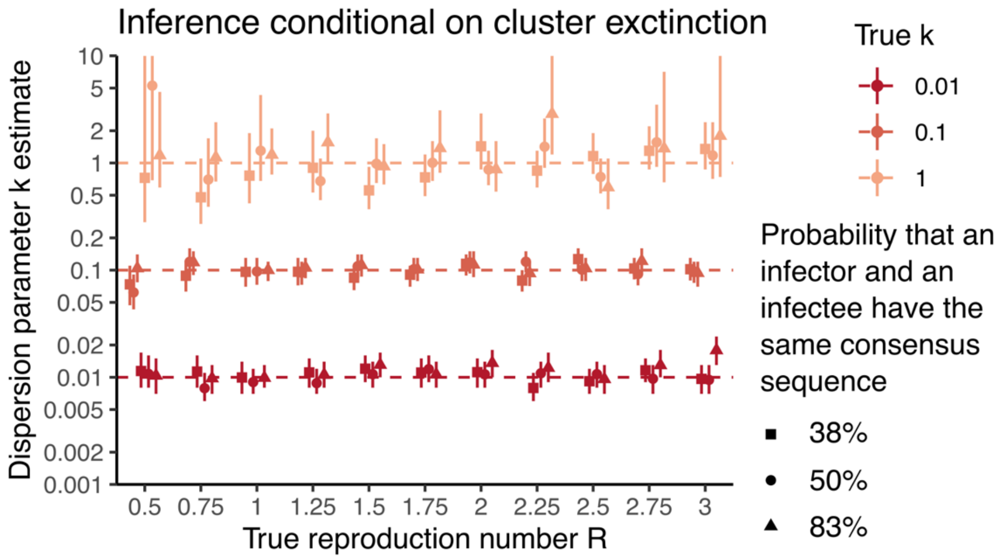


**Figure S21: Estimates of the dispersion parameter *k* using the size of clusters that got extinct.** Estimates are reported as a function of the true reproduction number *R* used to generate synthetic clusters. Point estimates correspond to maximum-likelihood estimates and vertical segments to 95% likelihood profile confidence intervals obtained from analyzing 1000 synthetic clusters of identical sequences.

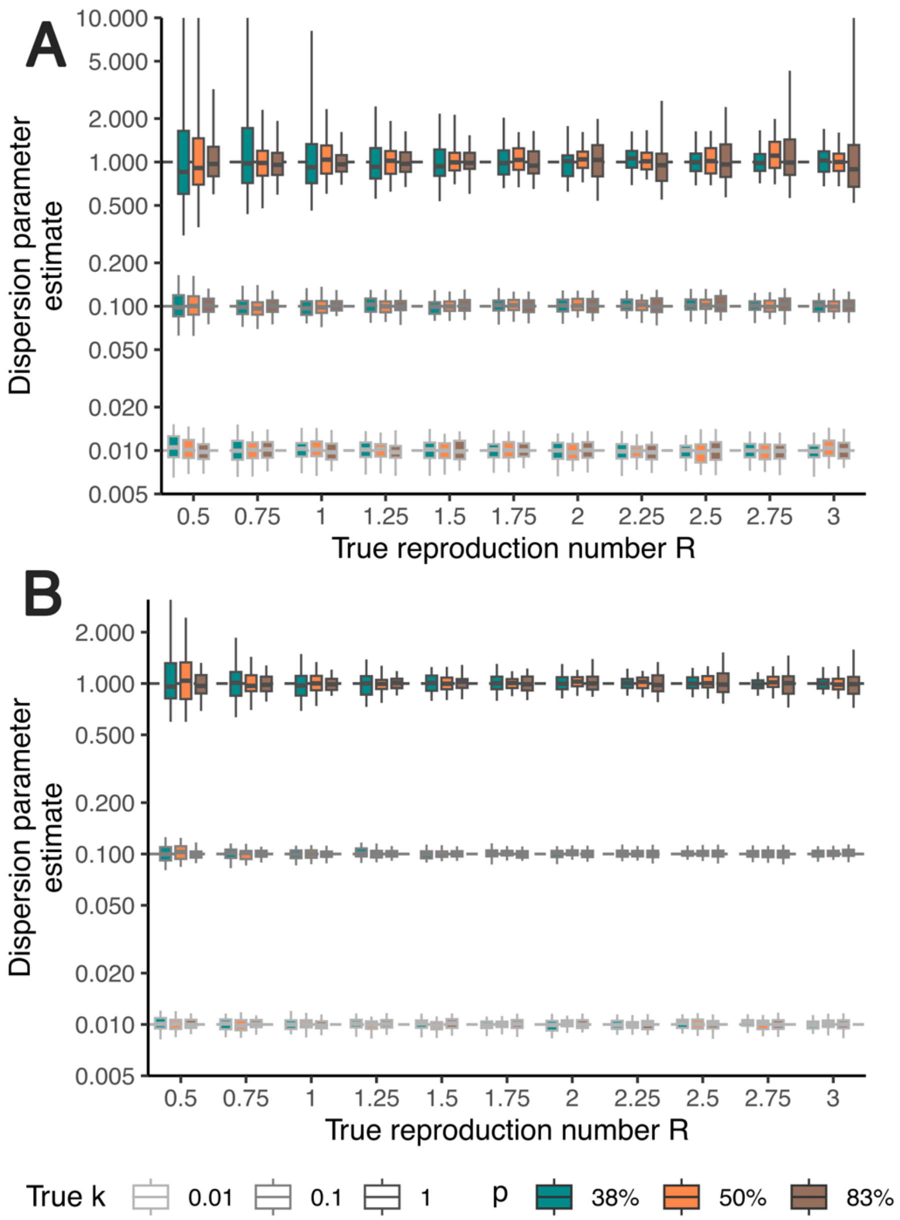


**Figure S22: Dispersion parameter estimates as a function of the true reproduction number when using the inference framework relying on cluster size distribution conditional on cluster extinction. A.** Using a dataset comprised of 1,000 clusters of identical sequences. **B.** Using a dataset comprised of 5,000 clusters of identical sequences. Each boxplot represents the distribution of *k* maximum likelihood estimates across 100 simulations (2.5%, 25%, 50%, 75% and 97.5% percentiles). We explored different values of the true dispersion parameter k (boxplot contour colours) and different values for the probability *p* that an infector and an infectee have the same consensus sequence (boxplot filling).

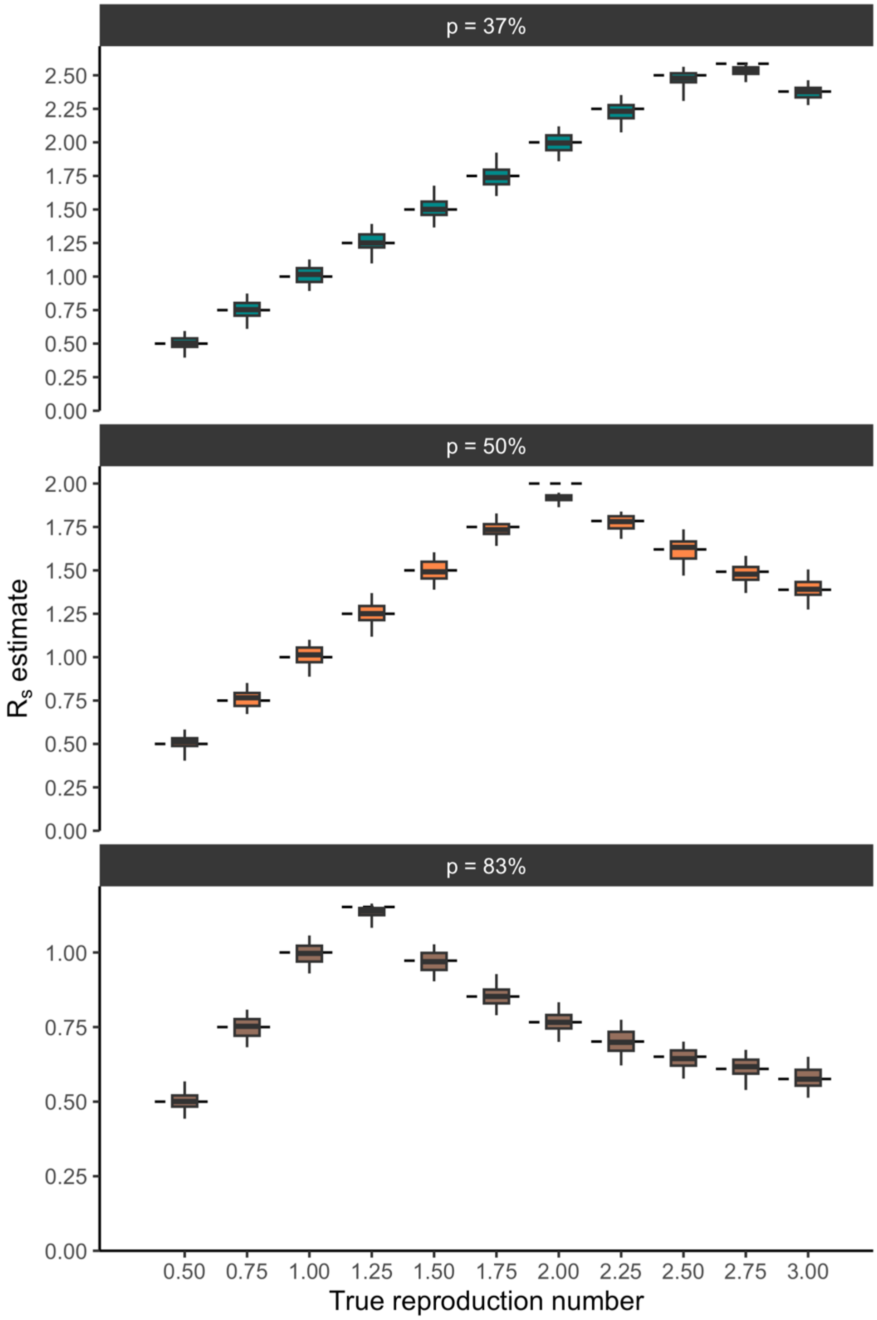


**Figure S23: Subcritical reproduction number $R_s$ estimates as a function of the true reproduction number when using the inference framework relying on cluster size distribution conditional on cluster extinction.** Each boxplot represents the distribution of $R_s$ maximum likelihood estimates across 100 simulations (2.5%, 25%, 50%, 75% and 97.5% percentiles). Results are displayed for a true dispersion parameter of 0.1 and running the inference on 1,000 clusters of identical sequences. Each panel corresponds to a different assumption regarding the probability p that an infector and an infectee have the same consensus sequence. The horizontal dashed segments correspond to the true value of $R_s$ (associated with the true reproduction number and the true dispersion parameter).

9. , GISAID - gisaid.org (9 February 2023).

10. ,*ncov-ingest: A pipeline that ingests SARS-CoV-2 (i.e. nCoV) data from GISAID and Genbank, transforms it, stores it on S3, and triggers Nextstrain nCoV rebuilds* (Github) (9 February 2023).

11. ,COVID-19 data portal (9 February 2023).

17. ,Nextstrain - Real-time tracking of measles virus evolution.
